# Meta-analysis of atopic dermatitis in 1,094,060 individuals identifies new risk loci, and sub-analysis characterizes the loci with disease severity and onset

**DOI:** 10.1101/2022.12.08.22283257

**Authors:** Anu Pasanen, Eeva Sliz, Laura Huilaja, FinnGen, Estonian Biobank Research Team, Ene Reimann, Reedik Mägi, Triin Laisk, Kaisa Tasanen, Johannes Kettunen

## Abstract

Atopic dermatitis (AD) is a common inflammatory skin disease highly attributable to genetic factors. Here, we report results from a genome-wide meta-analysis of AD in 37,541 cases and 1,056,519 controls with data from the FinnGen project, the Estonian Biobank, the UK Biobank, the EAGLE Consortium, and the BioBank Japan. We detected 77 independent AD-associated loci of which 10 were novel. The associated loci showed enrichment in various immune regulatory processes. We further performed subgroup analyses of mild and severe AD, and of early and late-onset AD, with data from the FinnGen project. 55 of the 79 tested variants in the associated loci showed larger effect estimates for severe than mild AD as determined through administered treatment. The age of onset, as determined by the first hospital visit with AD diagnosis, was lower in patients with particular AD-risk alleles. Our findings add to the knowledge of the genetic background of AD and may underlay the development of new therapeutic strategies.

---

Atopic dermatitis (AD) is a common chronic inflammatory skin disease with a lifetime prevalence of approximately 10-20%. AD can manifest at any age, but in most cases, AD symptoms start during the first year of life. In 70% of cases, AD onset occurs by the age of five years^1,2^. AD is characterized by dry, itchy skin and periodically occurring eczematous lesions. The pathophysiology of AD involves skin barrier dysfunction and impaired immune regulation of the innate and adaptive immune responses. Basic treatment of AD includes the use of emollients to protect and restore the skin barrier^3^. Medical therapies consist of topical corticosteroids or calcineurin inhibitors for patients with mild to moderate AD, and systemic approaches including cyclosporin, dubilumab, or janus-kinase (JAK)-inhibitors targeting the immune system in cases of severe AD^4^. Causes of AD are multifactorial including an interplay of genetic, epigenetic, and environmental factors^5,6^.

Genetic variation has a high contribution to AD susceptibility with heritability estimates of approximately 75% based on twin studies^7^. Previous genome-wide association studies (GWAS) have identified approximately 80 genetic risk loci that explain up to 30% of the variation in AD susceptibility^8–10^. The major predisposing genetic factors include loss-of-function (lof) variants in *FLG* gene encoding filaggrin protein. Filaggrin is essential in maintaining epidermal homeostasis, and *FLG* lof variants lead to skin barrier dysfunction that can lead to loss of hydration and make then skin prone to allergen and microbial entry^5^. Further risk loci contributing to epidermal strength and integrity include variants in desmocollin 1 (*DSC1*) and serpin family B member 7 (*SERPINB7*)^11^. Variants in multiple immune regulatory genes, such as interleukin-13 (*IL13*) and (*IL6R*), have been linked to immune dysregulation of AD^5^.

The occurrence of AD symptoms displays high heterogeneity during the course of the disease, which is reflected in specific aspects of the disease including the age of onset, disease severity and longitudinal trajectories of disease progression^12^. Previous studies have investigated some genetic variants in relation to AD severity, including variantions in *FLG*, and keratin 6A (*KRT6A*)^13,14^. *FLG* variants were associated with AD severity and early-onset AD in different populations^13^. Altogether, little is known about the associated variants in the context of AD severity or the age of onset.

In this study, we aimed to identify further genetic factors of AD in a meta-analysis of 1,094,060 individuals comprising 37,541 cases with AD and 1,056,519 healthy controls with data from FinnGen, UK Biobank, Estonian Biobank, EAGLE Consortium, and BioBank Japan. Another goal was to characterize the genetic background of AD disease subgroups that were classified as mild or severe based on the administered medical treatment, and as early or late-onset according to age during the first hospital treatment period related to AD diagnosis. We further aimed to investigate if particular risk genotypes were associated with the age of onset.

## Results

### Overview of the meta-analysis of AD

Meta-analysis of 1,094,060 individuals (37,541 cases; 1,056,519 controls) detected 77 associated loci with at least one variant associated with AD at *p*<5×10^−8^ (Figure 1, Table 1, Table S1). Majority of the identified associations were located in intronic or intergenic genomic regions, and genomic annotations of the associating variants were enriched in multiple categories (Figure S1). The distance between the associated loci was at least 1Mb with an exception of an association signal on chromosome one due to variants in the *FLG* locus that are known to span several megabases, and *DSC1* in chromosome 18 that was annotated as a causal gene in two loci with a rare missense variant rs200047736 and a noncoding variant rs1023133779 as lead variants. Our previous work indicates that the association signal linked to *DSC1* was driven by the missense variant, since the noncoding variant falls into the locus for which the association signal was attenuated upon conditioning with the missense variant^11^. Of the identified genome-wide significant loci, 10 were not previously associated with AD, as investigated in the NHGRI-EBI GWAS catalog^15^ with a +/-1Mb window around the meta-analysis lead variants or in a preprint presenting a multi-ethnic meta-analysis of AD^8^ (Table1, Table S2, Figure S2). Conditional association analysis of the novel AD-associated loci did not reveal secondary association signals (Figure S3).

**Table 1.**
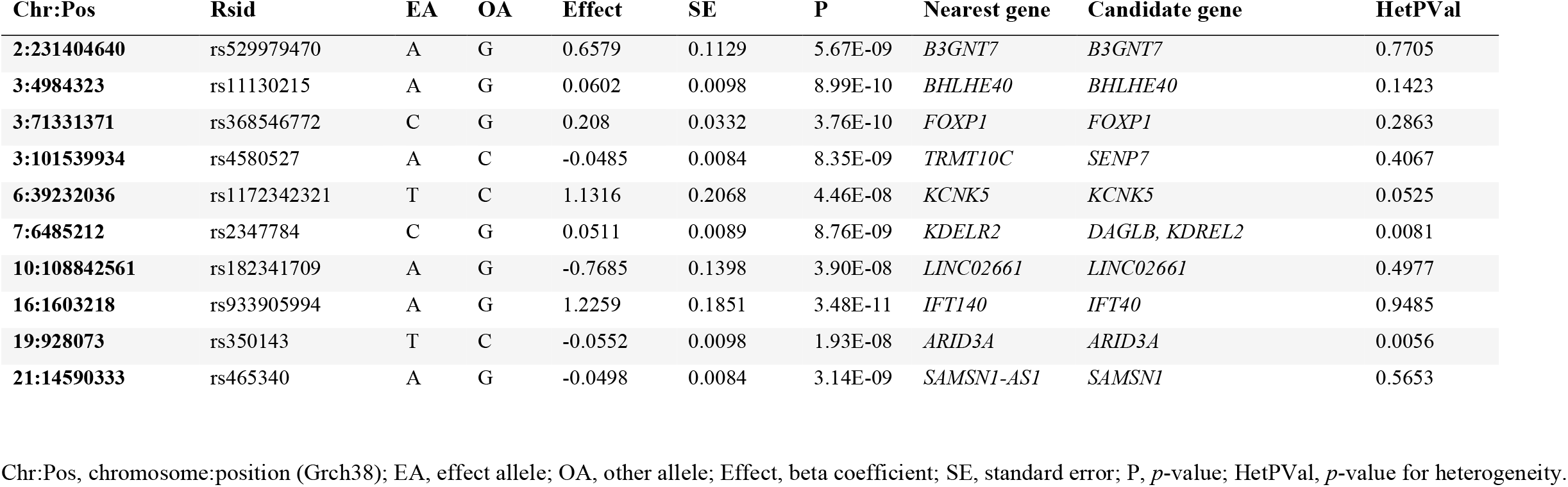
Novel loci associated with atopic dermatitis in the meta-analysis of 1,094,060 individuals from FinnGen, Estonian biobank, UK biobank, EAGLE consortium, and Biobank Japan. Variants represent individual associated loci with at least 1Mb distance to one another, and each locus has at least one variant with a genome-wide significant (*p*<5×10^−8^) association.

**Figure 1.**
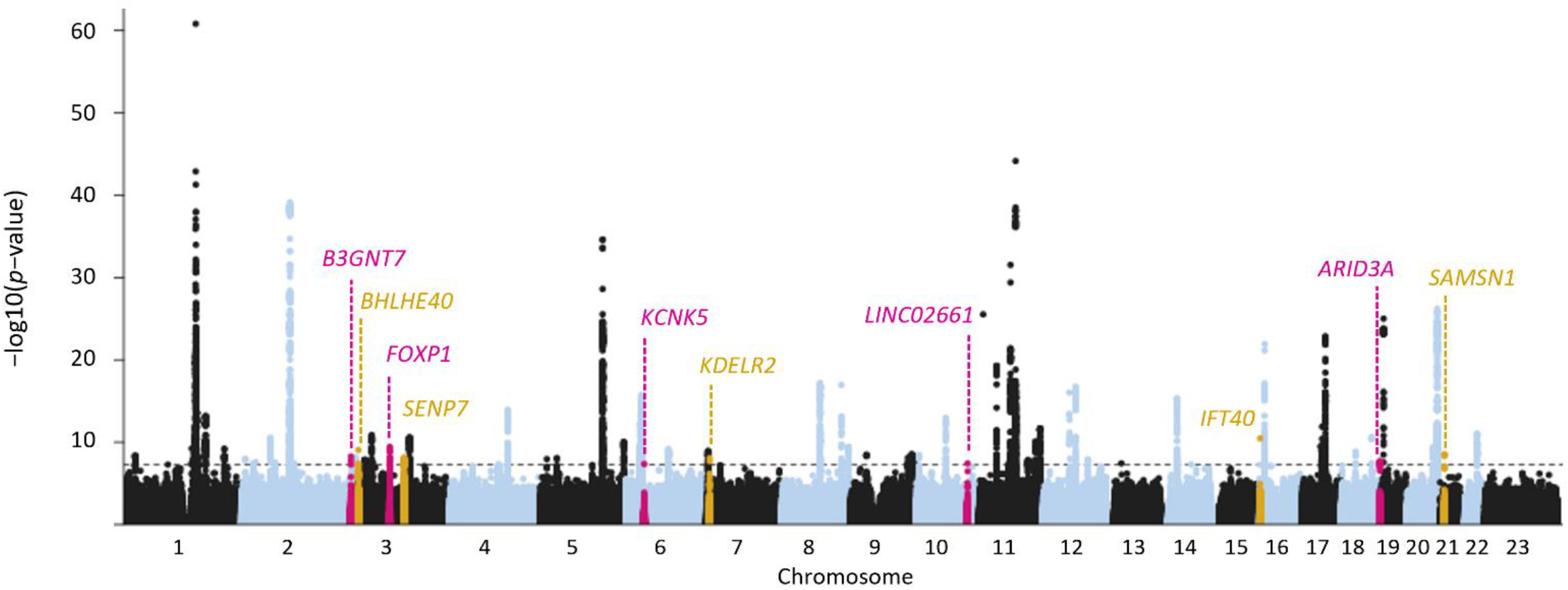
Manhattan plot of the loci associated with atopic dermatitis in the meta-analysis of 1,094,060 individuals (37,541 cases; 1,056,519 controls) from FinnGen, Estonian Biobank, UK Biobank, Biobank Japan, and European individuals of the EAGLE consortium. Variants are plotted on the x-axis according to their chromosomal positions, and the y-axis shows the corresponding association *p*-values at the -log10 scale. Dashed line denotes the threshold of genome-wide significance (*p*<5×10^−8^). The loci highlighted in pink or gold illustrate the ten novel AD-associated loci, defined as loci without previous AD-associations within a ±1 Mb window around the meta-analysis lead variant in the GWAS Catalog or in a recent meta-analysis of AD^8,15,58^.

65 of the 77 associated loci were genome-wide significant in the European-only meta-analysis (i.e., excluding BBJ, Table S3.) Of the the novel AD-associations, loci near *ARIDA3* and *KDELR2* had suggestive *p*-values in the European-only meta-analysis and reached genome-wide significance with the addition of BBJ, with all sub-population effects in the same direction. Of the previously known associations, seven showed suggestive or borderline-significant associations in the European-only meta-analysis. Loci near *NLRP10, GLB1*, and *INPP5D*, originally discovered in Asian populations, were not associated in the current meta-analysis with only European samples, which suggests population-specific associations at these loci.^16–19^.

We calculated the genome-wide inflation factor of the meta-analysis with linkage disequilibrium score regression (LDSC)^20^ (Figure S4). LDSC estimates indicated that the slightly inflated value of the factor (λ_GC_=1.1428) was mostly accounted by polygenic signal with the intercept being close to one (1.0472)^21^. LDSC-derived SNP heritability on a liability scale was 7.8% for the meta-analysis and 19.0% for the FinnGen sample alone. The heritability estimate based on only the European meta-analysis cohorts was similar (7.8%) compared to the whole meta-analysis. The estimates were in line with previous reports in which SNP heritabilities were estimated based on GWA data. Compared to our previous study of AD^11^, the heritability estimates increased approximately 2.4 and 4.7 percentage points in the meta-analysis and in the FinnGen sample alone, respectively.

Effect estimates for the genome-wide significant loci in individual study cohorts were largely concordant (Figure S5). Four relatively well-established AD risk loci, *IL18RAP, IL13, BACH2*, and *ZNF652* showed heterogeneity (*P*_adjusted_<0.05) (Table S1). In BBJ, the effect estimates were were remarkably similar with the European populations (Figure S5). *IL7R* was the only locus that showed a clear directional effect size difference between BJJ and the European cohorts. The association for the *IL7R* variant was not significant in BBJ (*P* = 0.32).

### Gene-set enrichment tests

MAGMA gene set enrichment of the genes annotated to the best loci indicated the involvement of several immune processes known to be important in AD pathophysiology (Table S4). As an example, pathways related to the regulation of lymphocyte activation, leukocyte mediated immunity, and interleukin-15 (*IL-15*) signalling, were well-represented. Pathways with fewer occurrences included for example Calcineurin-regulated NFAT-dependent transcription in lymphocytes, allograft rejection, and genes whose promoters are bound by FOXP3. Calcineurin inhibitors such as Cyclosporin are used to treat severe AD, and similar drugs are used to prevent allograft rejection after organ transplant^22^. We detected enrichment of AD-associated genes for the differentially expressed gene (DEG) sets in skin tissues among the 54 GTEx v8 tissues for both upregulated and 2-sided (up-and-down regulation) DEGs (Figure S6). The genes annotated as candidate genes in the novel AD risk loci were most overrepresented among the gene sets related to “cancer modules” and “immunologic_signatures”, tested against a background of all genes (Table S4)^23^. Among the traits in the GWAS catalog category, the novel AD associations were most overrepresented in cell type proportions in leukocytes and in inflammatory bowel disease (IBD).

### Colocalization analysis

We performed colocalization analysis to investigate if the risk variants influence AD by regulating mRNA or protein levels (table S6). Colocalization analysis indicated that matrix metallopeptidase locus, *MMP12*, affects AD susceptibility via regulation of matrix metalloproteinase levels (posterior probability, PP = 0.999) in the blood, and the AD-risk variants were associated with upregulated MMP12 level. Colocalization analysis at the level of gene expression indicated that many of the AD risk loci contribute to AD disease process via gene expression. As an example an AD-associated variant in the aquaporin 3 (*AQP3)* locus colocalized with the expression of *AQP7* and *AQP3* in the skin tissue. AD risk alleles were associated with increased expression of the *AQP7* (Table S3). Of the novel loci, we detected colocalization for *KDELR2* and *SENP7* in tissues including blood, and CD4 T-cells, CD8 T-cells, and monocytes.

### Survival analysis

Survival analysis detected a pattern of additive genetic effects, in which disease risk is proportional to the number of predisposing alleles in an individual, for the majority of the AD risk loci. Besides or in addition to additive genetic variance, allelic interactions within locus (dominance variance) and gene-gene interaction (epistasis) may contribute to the phenotypic variance of complex traits^24^. We detected evidence for non-additive genetic variance within the meta-analysis loci including *ZNF652, LRR32C, HLA-DQA1, TRIB1, OVOL1*, and *IL18RAP* (Figure S7). The variant in the *HLA-DQA1* locus showed the clearest deviation from the additive pattern. Human leukocyte antigen (*HLA*) genes encode cell surface proteins involved in various regulatory functions of the immune response. In line with our result, previous studies have found non-additive effects in several *HLA* genes in the context of immune system diseases that are closely related to AD, such as rheumatoid arthritis, psoriasis, and celiac disease^25^.

### Previous associations and potential functionality of novel risk loci

We screened the meta-analysis lead variants for trait associations in the FinnGen R7 data, in which genome-wide significant associations were only detected for AD (*ARID3A, BHLHE40*) (Figure 2). The number of suggestively associated traits (*p*<1e-05) was highest in diseases of the skin and subcutaneous tissue category, followed by diseases of the digestive system and diseases of the mucoskeletal system and connetive tissue -categories. In the GWAS catalog, variants in the novel AD risk loci were associated with multiple traits including blood cell counts or percentages for all loci except *LINC02661* (Table S2).

**Figure 2.**
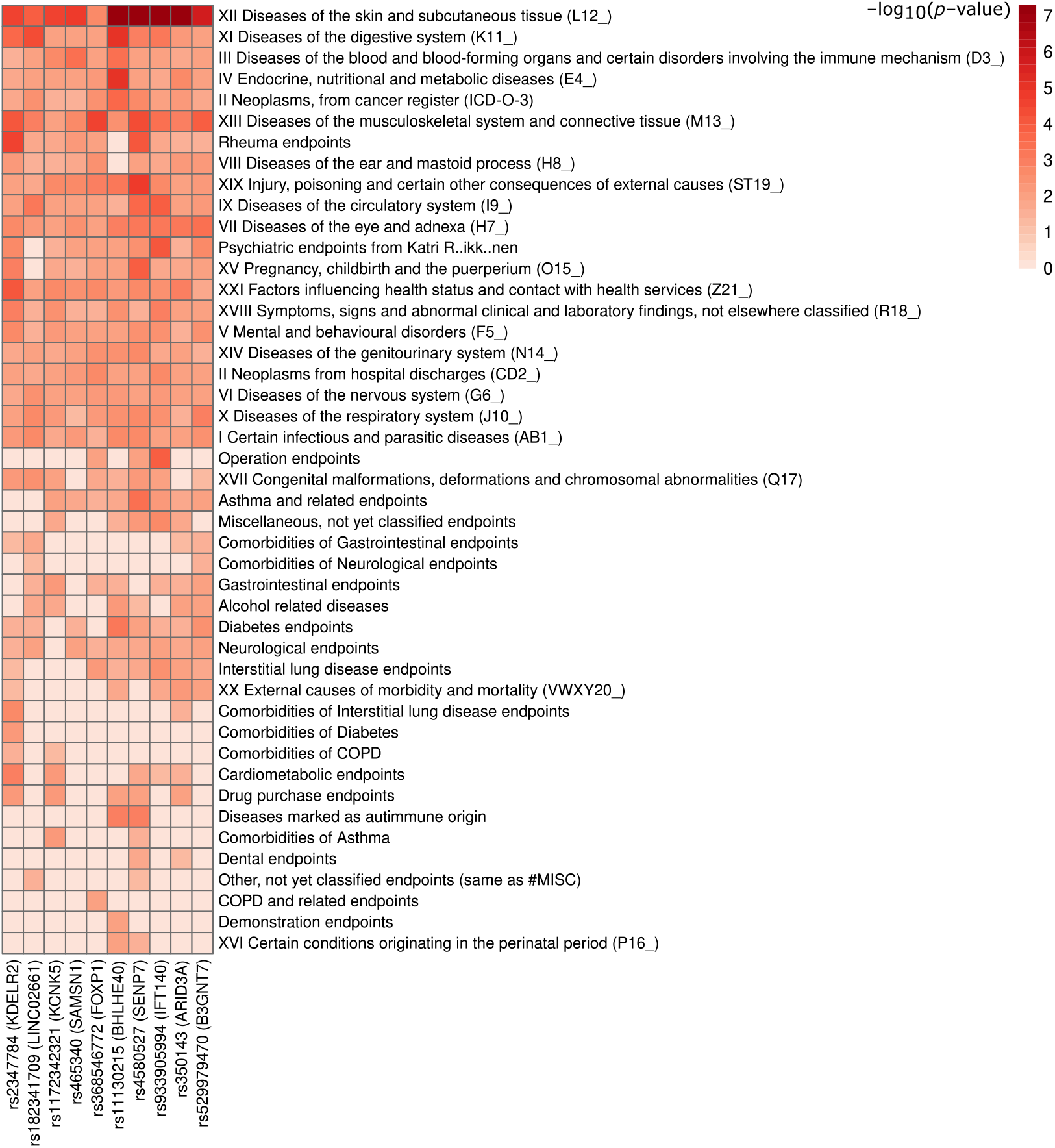
Associations of the novel AD risk loci accross the endpoint (phenotype) categories in the FinnGen freeze7. The FinnGen freeze7 comprizes approximately 300,000 individuals, and each endpoint category includes more than 3,000 phenotypes. *P*-values correspond to strongest within-category associations.

To assess the regulatory potential of the novel AD risk variants, we searched RegulomeDB^26^ for overlapping regulatory elements of the noncoding genome. 60% of the loci had at least one genome-wide significant variant that was ranked as potentially regulatory with probability >0.8 (Table S7). RegulomeDB also ranks variants with scores 1-6 with most to least support of binding and expression of a target gene. Of the meta-analysis lead variants, the index variants in *BHLHE40, ARID3A*, and *SAMSN1* had the strongest evidence for regulatory potential when we looked at the combination of both probability (>0.8) and ranking (>2) measures.

### Subgroup analysis of early-onset and late-onset AD

To analyze potential genetic differences related to the age of onset, we conducted FinnGen data-based analyses, in which we divided individuals into early-onset (<5 years, N = 728) and late-onset (>5years, N = 7,765) groups based on the age during the first hospital visit with AD-diagnosis. We detected a nominal effect size difference (*p*<0.05) for six of the AD meta-analysis risk loci; however, the effects were not consistently larger or smaller in either group (Table S8, Figure S8). Kaplan-Meier plots of time-to-AD onset stratified by genotype showed that some variants in the *FLG* locus, most clearly the deletion allele of the frameshift variant rs558269137, predispose to AD starting from early life (Figure 3). Other loci that were associated with high AD risk from childhood were *SERPINB7* and *TESPA1*, whereas for some of the AD risk loci, including *B3GNT7* and *LINC02661*, results suggested that the risk of AD starts to rise later in life (Figure 3). For *B3GNT7*, 18% of the cases with the AD risk-increasing genotypes (rs529979470 AA or AB) were classified as severe based on the medical treatment data (next paragraph), which was no different from 16% of severe AD cases within the non-risk genotype group (Fisher’s *p* = 0.696). For *LINC02661*, the majority (>90%) of the cases with the AD risk-increasing genotypes (rs182341709 G/G or A/G) were classified as mild.

**Figure 3.**
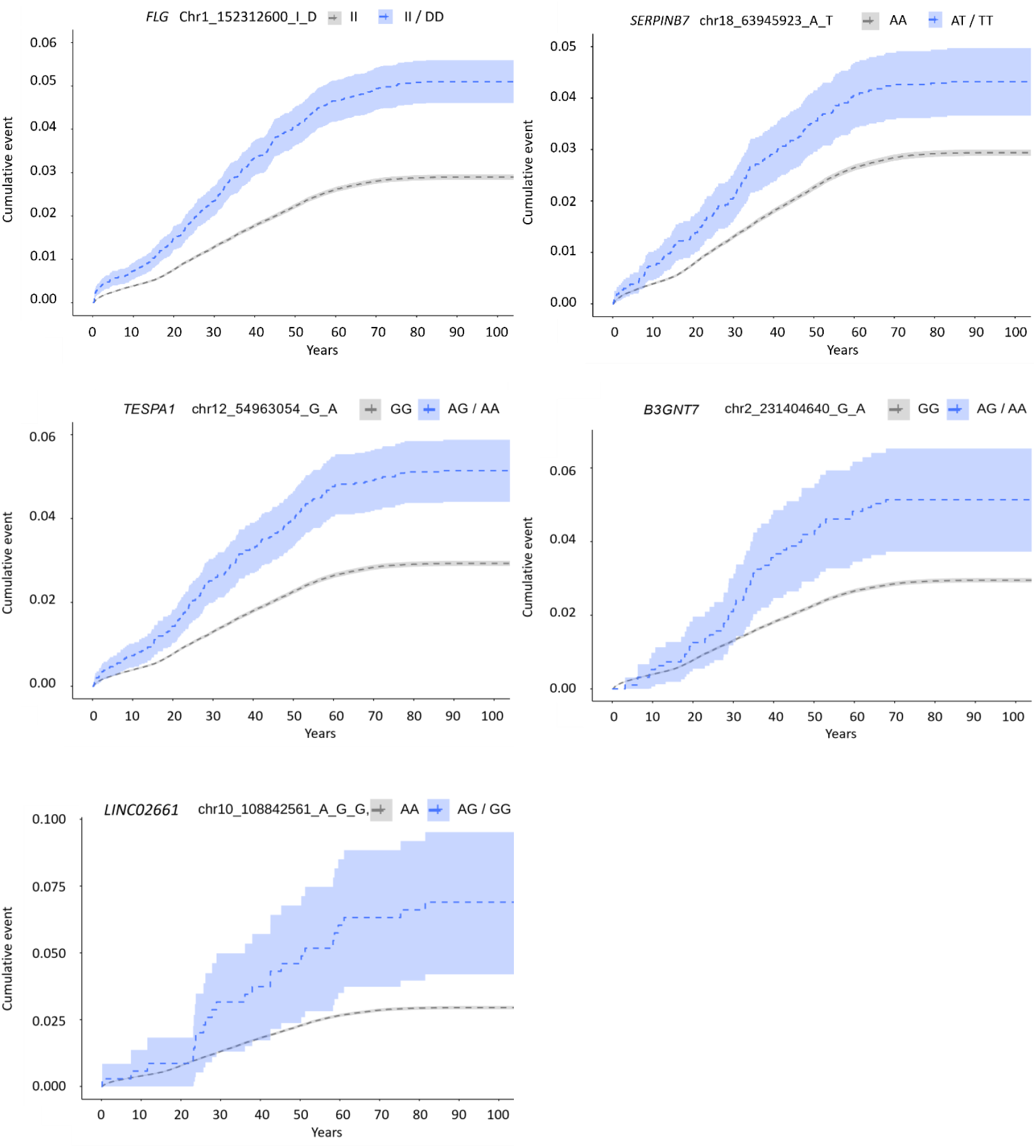
Kaplan-Meier plots of the survival analysis of AD onset per genotype in the FinnGen data. Disease onset is based on the first AD-related hospital visit. The risk loci associated with early-onset AD included *FLG, SERPINB7*, and *TESPA1*. The loci associated with late-onset AD included *B3GNT7* and *LINC02661*. To protect individual data, the rarer homozygous genotypes were combined with the heterozygous genotypes.

### Subgroup analysis of mild and severe AD

To investigate the genetic background of AD severity, we classified AD cases as mild or severe based on medical treatment information in the FinnGen data (Table S9, Figure S9). 70% of the effect sizes were larger in severe compared to mild AD, and we detected consistently larger effect sizes for six of the AD risk loci in severe *vs*. mild AD at *p*<0.05 (Figure4, Table S10). We further calculated polygenic risk score (PRS) based on the AD-associated meta-analysis loci, and observed a noticeable tendency toward larger effect sizes within severe compared to mild AD (Figure S10). The PRS was constructed for illustrative purposes of this analysis, and the conclusions are suggestive since the base and target populations had overlapping samples.

**Figure 4.**
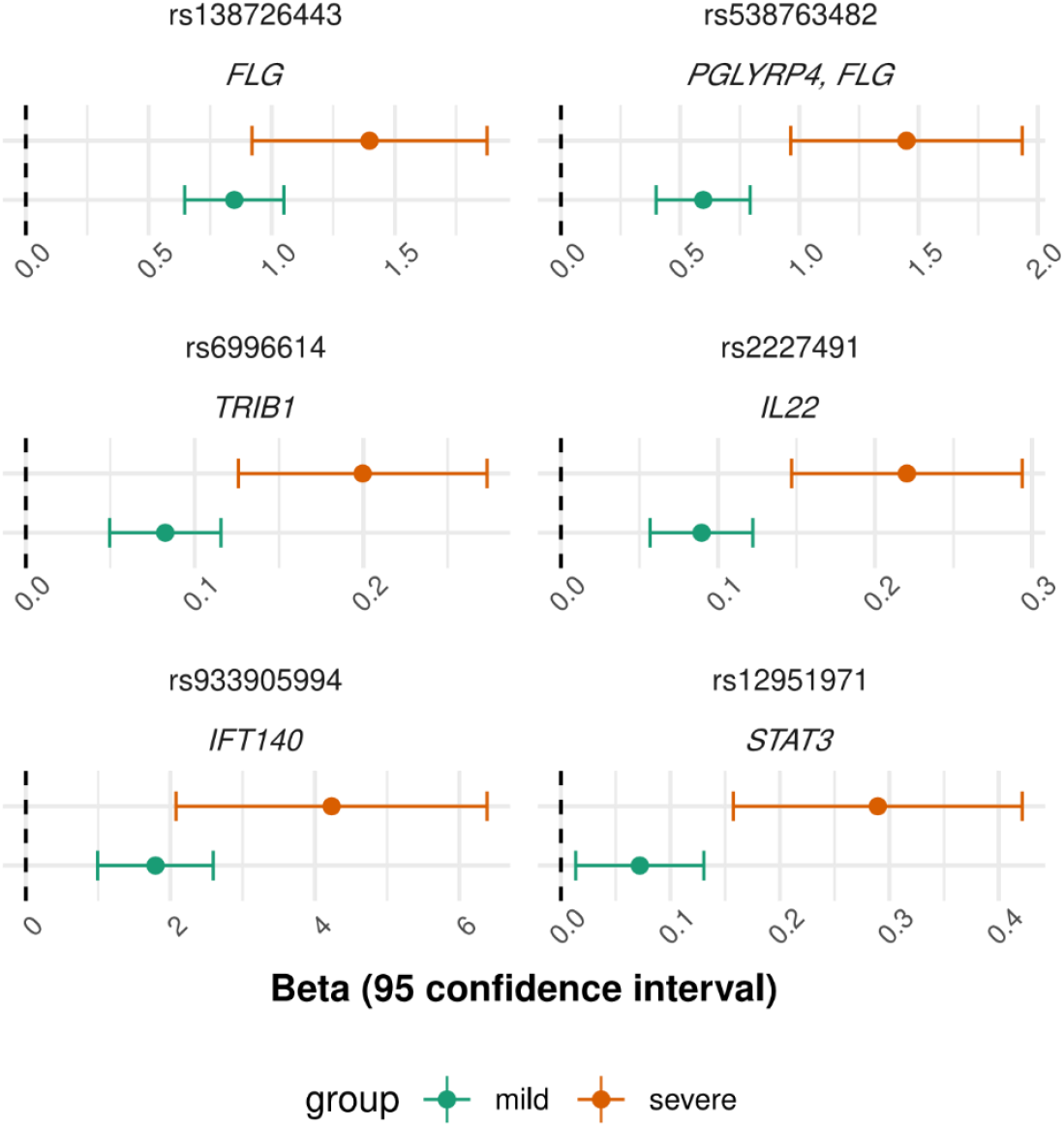
Effect size comparison of the AD-associated loci in mild and severe AD. Loci with *p*(Z_diff) <0.05 shown.

## Discussion

This genome-wide meta-analysis of 1,094,060 individuals with 37,541 AD cases and 1,056,519 controls detected 77 loci that were independently associated with AD. Genomic analyses with the genome-wide association landscape highlighted the involvement of T cell-mediated and various other immunological pathways in AD pathophysiology, accentuated the close connection among AD and other inflammatory diseases, and emphasized the importance of the skin and inflammatory cells as the most relevant tissue and cell types in AD disease processes. Besides previously known pathways of AD, the pathway analysis indicated the involvement of genes whose promoters are bound by FOXP3. *FOXP3* encodes a transcription factor forkhead box P3 expressed in natural T-regulatory cells (Tregs) and in a subset of adaptive Tregs, where FOXP3 can act as a transcriptional activator or repressor of the bound genes. Tregs play important roles in immune responses and homeostasis of the skin, and FOXP3 is vital to Treg function. Mutations in *FOXP3* have been associated with immune dysfunction and eczematous dermatitis that resembles severe AD^27–29^. Our results suggest that FOXP3-mediated signalling of the AD-associated genes leads to altered function of Tregs in the skin of AD patients. However, results from studies of altered Treg frequency in AD patients remain elusive, and the exact role of FOXP3-regulated AD-risk genes and their relation to Treg function in the skin tissue of AD patients warrants further study.

Colocalization analysis revealed that many of the AD risk loci may contribute to AD by regulating gene or protein expression. At protein level, *MMP12* likely contributes to AD disease process via regulating MMP12 expression. *MMP12* is a member of the matrix metalloproteinase cluster involved in the degrading of the extracellular matrix under normal physiological conditions and in disease processes. Recent meta-analysis of AD^8^ detected an association in the locus, but the potential role of *MMP12* in AD disease process in not known. *MMP12* was previously associated with diseases including emphysema and chronic obstructive pulmonary disease (COPD), and the meta-analysis lead variant showed associations with similar phenotypes in the GWAS catalog. Elevated levels of MMP12 were previously associated with ultraviolet A1 (UVA1) induced damage of the skin^30^, and molecular biological studies have linked MMP12 with T cell-mediated immune response in the skin and the gut of dermatitis herpediformis patients^31^, and as a molecule of the blood signature of AD^32^. Our results suggest that the *MMP12* locus affects AD via elevated MMP12 levels under inflammatory conditions.

Variants in loci including *AQP3, SENP7*, and *KDELR2* were associated with gene expression. *AQP3* locus was only recently found associated with AD^8^, and associations for *SENP7* and *KDELR2* were discovered in the current study. AD-associated variant in the aquaporin 3 (*AQP3)* locus colocalized with the expression of *AQP7* and *AQP3* in the skin tissue. *AQP3* and *AQP7* are broadly expressed water- and glycerol channels in cell membranes, and both molecules were linked to keratinocyte function and inflammatory skin disease in previous molecular biological investigations^33–36.^ The AD-associated allele was linked with increased expression of *AQP7*, whereas increased expression of *AQP3* was previously detected in eczema compared to healthy skin^33^. It was further indicated that increased *AQP* expression leads to loss of hydration, a process that could compromise epithelial integrity and barrier function in AD patients. Thus, both *AQP3* ad *AQP7* represent fascinating candidates for further molecular biological investigations related to AD. *SENP7* (SUMO specific peptidase 7) and *KDELR2* (KDEL endoplasmic reticulum protein retention receptor 2) variants colocalized with expression of the respective genes in various blood cell types. Interestingly, *SENP7* variants colocalized with *SENP7* expression in neutrophils and lymphocytes, both involved in immune systems’ defenfe against external agents. Interestingly, *SENP7* was previously linked to allergic disease^37^. *KDERL2* plays a role in protein trafficking in endoplastic reticulum, and *KDELR2* variant colocalized with *KDELR2* gene expression in blood, monocytes, and CD4 and CD8 T-cells.

Ten of the identified loci were not previously associated with AD at the time of this investigation. Variant associations in the new AD risk loci point to several promising candidate genes that were prioritized based on known biology, functional or statistical annotations, and QTL colocalization analysis. Previous associations of the novel AD risk loci included mainly other inflammatory diseases and cell type counts or proportions of the inflammatory cell types. Interestingly, some of the genes had associations with diseases of the digestive system, such as IBD, and the new AD candidate genes together were enriched for IBD associations. AD and IBD have been shown to correlate^38,39^, and our findings provide genetic links between the two clinically different diseases. Many of the novel AD-associated loci had putative regulatory functions that may play a role in AD pathophysiology. The meta-analysis lead variants in *BHLHE40, ARID3A*, and *SAMSN1* showed high likelihood of being regulatory. *BHLHE40*, Basic Helix-Loop-Helix Family Member E40, was recently recognized as a central regulator of inflammation^40,41^. The adjacent lead SNP rs11130215 in the *BHLHE40* 3’UTR binds to multiple transcription factors in the blood, including RUNX family transcription factor 3 (*RUNX3*) that was associated with AD in previous GWA studies. The lead variant rs350143 in AT-rich interaction domain 3A, *ARID3A*, had binding sites to several transcription factors, including argonaute RISC catalytic component 2 (*AGO2*) that was only recently discovered to be associated with AD^8^. *ARID3A* is a B-cell restricted transcription regulator of immunoglobulin heavy chain, and the locus had some recent associations with AD-related traits including autoimmune and inflammatory disease and immune dysregulation preceding asthma^42,43^, whereas *AGO2* has a central role in the biogenesis of miRNAs. *AGO2* has been shown to participate in inflammation-related wound healing and was recently found to be upregulated in keratinocytes of psoriasis patients^44,45^. SH3 domain and nuclear localization signals 1, *SAMSN1*, is a negative regulator of B-cell activation that is up-regulated in peripheral blood B-cells by IL4 and IL13 and by CD40 stimulation. Interestingly, differential expression of SAM domain was previously detected in the transcriptomic signatures of AD patients^46^.

In the sub-analysis of the age of AD onset, we detected loci that confer a high AD risk starting from childhood, and loci that were associated with AD later in life. Besides *FLG* risk alleles with previous association with early-onset AD, we detected that risk genotypes in *SERPINB7* and *TESPA1* were associated with AD risk starting early in life. We speculate that rather than some risk loci predisposing to late-onset AD, it is more likely that the observed patterns were driven by a low and delayed need for specialist care due to mild AD.

We further found support for differing genetic architecture in mild *vs*. severe AD, when we assigned AD patients into severity groups based on administered treatments. We found consistently larger effect sizes and PRS estimates for severe compared to mild AD, implying that the risk of severe AD is likely more extensively affected by genetic factors and more accurately predicted by the PRS. Interestingly, a recent study found that PRS estimates correlated with AD severity defined by e.g. Rajka Langeland score and Eczema Area and Severity Index (EASI)^47^. Unfortunately, in our current study based on nationwide health registeries it was not possible to obtain more detailed clinical data.

Altogether, the genome-wide findings of the meta-analysis fit well with the current understanding of AD, and strengthen the view of relevant tissues, cell types, and pathways involved in AD pathophysiology. Through gene-set analysis, genomic annotations, and colocalization, this study highlighted additional biological routes that connect genetic variants to AD pathophysiology. These in silico functional analyses all implicated immune cells and the skin as tissue types of interest, and emphasized the role of immunological processes, providing genetic support to the current understanding of AD pathophysiology. The discovery of new loci and candidate genes enhances our understanding of AD risk and provides new opportunities to gain insight into AD etiology and develop new treatments.

## Methods

### Study Populations

The current meta-analysis of AD was conducted with data from 1,094,060 individuals (37,541 cases; 1,056,519 controls) originating from FinnGen research project, Estonian Biobank, UK Biobank, EAGLE Consortium, and Biobank Japan. The **Finngen research project** (www.finngen.fi)^48^, launched in 2017, combines genome data with digital health care information via national personal identification numbers. The final resource will contain data from 500,000 participants. The current GWAS was conducted with data from 289,072 Finnish adults comprising 10,277 AD cases and 278,795 controls from FinnGen Preparatory Phase Data Freeze 7. **The Estonian Biobank** is a population-based collection of adult volunteers that currently has genome data and health care information from 200,000 Estonian residents. The present GWAS analyzed data from 136,724 participants comprising 11,187 AD cases and 125,537 controls. **The UK Biobank** (UKBB) brings together genetic data and health care information from 500,000 study participants from UK, aged 40-69 years. The current GWAS analyzed data from 425,393 individuals comprising 2,904 cases and 412,489 controls, as reported in the Pan-UKB project^49^. **The EAGLE Consortium** (EArly Genetics and Lifecourse Epidemiology; https://www.eagle-consortium.org/)^50^ is a collaborative consortium of pregnancy and birth cohorts that seeks to study the genetic basis of various phenotypes from antenatal and early life to adolescence. The present AD GWAS was based on 40,835 individuals (10,788 cases; 30,047 controls) of European ancestry^51^. **The BioBank Japan** (BBJ)^52^, launched in 2003, comprises genome data and clinical information from 260,000 patients that originate from 12 medical institutions across Japan, and represent approximately 50 complex disease phenotypes. For the current study, data from 212,036 study subjects (2,385 cases; 209,651) were investigated. The data was downloaded from Japanese ENcyclopedia of GEnetic associations by Riken (Jenger; http://jenger.riken.jp/en/).

### FinnGen ethics statement

Patients and control subjects in FinnGen provided informed consent for biobank research, based on the Finnish Biobank Act. Alternatively, separate research cohorts, collected prior the Finnish Biobank Act came into effect (in September 2013) and start of FinnGen (August 2017), were collected based on study-specific consents and later transferred to the Finnish biobanks after approval by Fimea (Finnish Medicines Agency), the National Supervisory Authority for Welfare and Health. Recruitment protocols followed the biobank protocols approved by Fimea. The Coordinating Ethics Committee of the Hospital District of Helsinki and Uusimaa (HUS) statement number for the FinnGen study is Nr HUS/990/2017.

The FinnGen study is approved by Finnish Institute for Health and Welfare (permit numbers: THL/2031/6.02.00/2017, THL/1101/5.05.00/2017, THL/341/6.02.00/2018, THL/2222/6.02.00/2018, THL/283/6.02.00/2019, THL/1721/5.05.00/2019, THL/1524/5.05.00/2020, and THL/2364/14.02/2020), Digital and population data service agency (permit numbers: VRK43431/2017-3, VRK/6909/2018-3, VRK/4415/2019-3), the Social Insurance Institution (permit numbers: KELA 58/522/2017, KELA 131/522/2018, KELA 70/522/2019, KELA 98/522/2019, KELA 138/522/2019, KELA 2/522/2020, KELA 16/522/2020, Findata THL/2364/14.02/2020 and Statistics Finland (permit numbers: TK-53-1041-17 and TK/143/07.03.00/2020 (earlier TK-53-90-20). The Biobank Access Decisions for FinnGen samples and data utilized in FinnGen Data Freeze 7 include: THL Biobank BB2017_55, BB2017_111, BB2018_19, BB_2018_34, BB_2018_67, BB2018_71, BB2019_7, BB2019_8, BB2019_26, BB2020_1, Finnish Red Cross Blood Service Biobank 7.12.2017, Helsinki Biobank HUS/359/2017, Auria Biobank AB17-5154 and amendment #1 (August 17 2020), Biobank Borealis of Northern Finland_2017_1013, Biobank of Eastern Finland 1186/2018 and amendment 22 § /2020, Finnish Clinical Biobank Tampere MH0004 and amendments (21.02.2020 & 06.10.2020), Central Finland Biobank 1-2017, and Terveystalo Biobank STB 2018001.

### Phenotype definitions

In FinnGen, AD cases were defined based on International Classification of Diseases (ICD) codes with a requirement for L20 (ICD-10), 6918 (ICD-9; 6918X excluded) or 691 (ICD-8) entry in the Hospital Discharge Registry, cause of death registry, or Finnish Social Insurance Institution (KELA) registry. Individuals without a record of the mentioned ICD codes were deemed controls. Similarly, in the Estonian Biobank and the UKBB, study subjects with the L20 code were classified as cases, and participants without a record of the L20 were treated as controls. In the EAGLE Consortium, AD case/control definitions were based on questionnaires of self-reported or doctor-diagnosed AD. In the BBJ, study participants of Japanese origin were defined as cases based on diagnosis by physician the criteria of Hanifin and Rajka^18,53^. the control population comprised participants without AD or AD-related disease, such as asthma, and without autoimmune disease such as rheumatoid arthritis.

### Genotyping, imputation, and quality control

FinnGen samples were genotyped with Illumina and Affymetrix chip arrays (Illumina Inc., San Diego, and Thermo Fisher Scientific, Santa Clara, CA, USA). Samples with ambiguous sex, non-Finnish ancestry, high missingness (>5%), and excess heterozygosity (+-4SD) were excluded. Variant quality control (QC) entailed excluding variants with high missingness (>2%), minor allele count, MAC, <3, and deviation from HWE (*p*<1e-6). Genotype imputation was performed against a Finnish population specific SISu reference panel with Beagle 4.1. Variants with imputation INFO<0.7 were excluded^54^.

Samples from the Estonian Biobank were genotyped with Illumina arrays (GSAMD-24v1, GSAMD-24v2, ESTchip-1_GSAv2-MD, and ESTchip-2_GSAv3-MD) in the Core Genotyping Lab of Institute of Genomics, University of Tartu, Estonia. Sample QC entailed excluding individuals with high missingness (>5%) or ambiguous sex. Variants with low call rate (<95%), minor allele frequency (MAF) <1% or deviation from HWE (*p*<1E-4) were excluded. Genotype imputation was done with Beagle v.28Sep18.793 against and Estonian-specific reference^55^. Indels were removed prior to imputation.

In the UKBB, imputed variants with INFO score > 0.8 from the UKBB version 3 were analyzed^49^.

AD summary statistics from the EAGLE consortium were based on 22 European cohorts. Genotype imputation with 1000 Genomes Project Phase 1 reference panel was conducted separately on individual cohorts. QC entailed excluding variants with MAF<1% or poor imputation quality score (Rsq<0.3 for variants imputed in MACH; proper info<0.4 for variants imputed with IMPUTE)^51^.

BBJ samples were genotyped with Illumina HumanOmniExpressExome BeadChip or a combination of the Illumina HumanOmniExpress and HumanExome BeadChips. Sample QC entailed excluding samples with call rate <0.98 and individuals that were not of East Asian ancestry. In genotype QC, variants with call rate < 99%; deviation from HWE (*p*<1E-6) or presence of less than five heterozygotes were excluded. Imputation was performed against the 1000 Genomes Phase 3 (version 5) reference panel with minimac3, version 2.0.1. Variants with an imputation quality metric Rsq <0.7 were excluded^56^.

### GWAS

GWAS of the FinnGen samples was done with REGENIE two-step regression model implemented in an automated pipeline at FinnGen environment. Age, sex, genotyping batch, and 10 top principal components were used as covariates. MAC was set to 5 for cases and controls to be retained in the analysis.

Samples from the Estonian Biobank, the UKBB, and the BBJ were analyzed with SAIGE (Scalable and Accurate Implementation of Generalized) software. In the Estonian Biobank, association models were adjusted for age, sex, and the leading 10 principal components. Variants with MAC of at least five were included in the analysis. In the UKBB, GWAS covariates comprized age, sex, age*sex, age2, age2*sex, and the first 10 principal components.

Variants with MAC>20 were retained in the analysis. GWAS of the BBJ incorporated age, sex, and the first five principal components as covariates, and variants with MAC<10 were excluded.

In the EAGLE Consortium, GWA analyses were done separately on each of the 22 cohorts with European study participants. GWA analyses were conducted with logistic regression implemented in several software, after which the studies were meta-analyzed with fixed-effect meta-analysis using GWAMA^10^.

### Meta-analysis

METAL was used to conduct fixed-effect inverse variance-weighted meta-analysis of the summary data obtained from the FinnGen, the Estonian Biobank, the UKBB, the EAGLE Corsortium, and the BBJ^57^. We also meta-analyzed cohorts of only European origin. We report associating loci based on at least two of the five meta-analysis populations. Genome-wide inflation factor of the meta-analysis was calculated with linkage disequilibrium score regression (LDSC)^21^. We considered the standard genome-wide significance level (*p*<5E-8) as a threshold of significant association.

### Characterization of the associating loci

Associated loci were defined as genomic regions within +/-1Mb window around the lead AD-associated variant. Thus, all associated loci were at least 1Mb apart and harbored at least one variant with *p*<5E-8. The locus was defined as novel if there were no significant AD-associations reported in the NHGRI-EBI Catalog of human genome-wide association studies^15,58^ or in a recent meta-analysis of AD^8^ within the +/-1Mb window around the index variant. We further performed conditional association tests with Genome-wide Complex Trait Analysis (GCTA) software package^59^, to dissect the risk loci for secondary association signals in the novel loci. In the conditional analyses, FinnGen was used as a reference sample to estimate linkage disequilibrium (LD) corrections. The associations were conditioned for the loci-specific lead variants with the exception of two loci on chromosomes 3 and 7, for which the genotype information of the lead variants was not available in FinnGen; in the case of these two loci, the associations were conditioned for the 2^nd^ most significant variants (chr3:71329215 C/CT and chr7:6462736 T/C, respectively).

To characterize associated loci and prioritize candidate genes within the novel loci, we gathered evidence from various approaches, as described below. We investigated gene and protein functions in GenBank and Uniprot^60,61^, and performed a thorough literature search to obtain knowledge of the previous studies concerning the genes of interest. We used DEPICT (Data-driven Expression Prioritized Integration for Complex Trait)^62^ implemented in CTG-VL (Complex-Traits Genetics Virtual Lab)^63^ to provide statistical support for the most likely causal gene. DEPICT leverages co-regulation of gene expression together with previously annotated gene sets in the gene prioritization process^62^. For the known loci, we report candidate genes mentioned in previous studies. We investigated associated traits in the FinnGen R7 data. We used FUMA (Functional Mapping and Annotation of Genome-Wide Association Studies), an integrative web-based application that leverages information from various biological data repositories in functional annotation of GWAS results^23^, to perform functional mapping, prioritize genes for gene-based enrichment test, and assess tissue-specificity by testing enrichment of differentially expressed gene (DEG) sets in Genotype-Tissue Expression (GTEx) v8 tissue types^64^. Enrichement of DEG sets is calculated in one tissue compared to all other tissue types within a given source of tissues^23^. FUMA implements MAGMA^65^ in gene-based analysis and gene-set enrichment analysis that is performed for GWAS summary data using curated gene sets and GO terms from MsigDB. In addition, the novel candidate genes were tested for enrichment against various gene sets with hypergeometric tests in the GENE2FUNC process^23^. We further performed colocalization analysis with HyprColoc^66^, a Bayesian method for finding shared genetic associations among complex traits in a particular genomic region, to assess if the loci (index variant +/-500 kb) contribute to AD risk by affecting gene or protein expression. We used cis-eQTLs (FDR>0.5) from, eQTLGen^67^ and eQTL Catalogue^68^, and protein level data assayed in plasma^69^. Of the GTEx data in the eQTLCatalogue, we included GTEx v8 and LCLs from an earlier release. We report colocalization results with a posterior probability (PP)>0.6. ^24^We performed survival analysis with R’s^70^ survival package to assess inheritance models of the meta-analysis risk loci.

### SNP-based heritability

We estimated SNP-based heritability on a liability scale for the meta-analyzed sample and the Finngen GWAS with LDSC^21^. Because LDSC uses European data as a reference for LD, we also estimated heritability from meta-analysis based on European samples only (BBJ excluded). We used a population prevalence of 15 %^11^ and sample prevalences of 3.40 % for the whole meta-analysed sample, 3.99 % for the meta-analyzed sample excluding BBJ, and 3.56 % for the FinnGen sample.

### Subgroup analysis of early-onset and late-onset AD

To assess the genetic background of early-onset and late-onset AD, we used FinnGen data and Hospital Discharge Registry data that contain information about all inpatient and outpatient visits in Finnish hospitals, including duration and diagnosis. We defined the early-onset and the late-onset AD based on hospital treatment periods related to AD (main or side diagnosis in the Hospital Discharge Register) as a dichotomous outcome starting before or after five years of age, and performed GWAS on both classifications with SAIGE using the population without AD as a control group. We then compared the effect size estimates of the AD risk loci from the current meta-analysis in the early and late-onset groups. We investigated the role of AD risk loci in early and late-onset AD with survival analysis of time-to-AD onset stratified by risk variant genotypes. To protect individual data according to regulations of the FinnGen project, we combined rarer homozygous genotypes with heterozygous genotypes when the locus under investigation had low MAC.

### Subgroup analysis of mild and severe AD

To investigate the genetic background of AD severity, we classified AD cases in the FinnGen data as mild or severe based on the medical therapy they had received. Medication data was derived from the Social Insurance Institution of Finland (KELA), which includes data on all reimbursed drug purchases from 1995 onwards. Classifications by Anatomical Therapeutic Chemical (ATC) codes were used in the analysis. Patients without medical treatment or with only topical corticosteroids (ATC codes: D07AD01, D07AC01, D07BC01, D07CC01, D07AC13, D07AC03, D07AC14, D07AB02, D07BB04, D07AB01, D07AB08, D07AA02, D07BA04) were defined as mild, whereas patients treated with topical calcineurin inhibitors (H02AB06, H02AB04) or systemic treatment (L04AX01, L01BA01, L04AD01, D11AH05) were classified severe. Patients with only systemic corticosteroid treatment (H02AB06, H02AB04) were excluded, since systemic steroids are widely used to treat AD patients with symptoms ranging from mild to severe. The groups have no overlapping samples, and the group memberships were assigned starting from the most severe AD. We used SAIGE to perform GWAS for the mild and severe groups of AD using individuals without AD as controls. We subsequently extracted the significant AD-associated loci based on the current meta-analysis and compared the effect size estimates of the mild and severe GWASs. We further constructed a polygenic risk score (PRS) based on the AD-associated meta-analysis loci, and calculated the PRS for the mild and severe AD cases in the FinnGen data with Plink1.9^71^. The PRS was constructed for illustrative purposes and it is only descriptive, because the base and target populations had overlapping samples

## Data Availability

Summary statistics of the meta-analysis results will be deposited in an appropriate data repository after publication of the manuscript.

## Estonian Biobank Research Team

Mari Nelis

Lili Milani

Tõnu Esko

Andres Metspalu

Georgi Hudjashov

## Acknowledgements

We want to acknowledge the participants and investigators of the FinnGen study. The FinnGen project is funded by two grants from Business Finland (HUS 4685/31/2016 and UH 4386/31/2016) and the following industry partners: AbbVie Inc., AstraZeneca UK Ltd, Biogen MA Inc., Bristol Myers Squibb (and Celgene Corporation & Celgene International II Sàrl), Genentech Inc., Merck Sharp & Dohme Corp, Pfizer Inc., GlaxoSmithKline Intellectual Property Development Ltd., Sanofi US Services Inc., Maze Therapeutics Inc., Janssen Biotech Inc, Novartis AG, and Boehringer Ingelheim. The following biobanks are acknowledged for delivering biobank samples to FinnGen: Auria Biobank (www.auria.fi/biopankki), THL Biobank (www.thl.fi/biobank), Helsinki Biobank (www.helsinginbiopankki.fi), Biobank Borealis of Northern Finland (https://www.ppshp.fi/Tutkimus-ja-opetus/Biopankki/Pages/Biobank-Borealis-briefly-in-English.aspx), Finnish Clinical Biobank Tampere (www.tays.fi/en-US/Research_and_development/Finnish_Clinical_Biobank_Tampere), Biobank of Eastern Finland (www.ita-suomenbiopankki.fi/en), Central Finland Biobank (www.ksshp.fi/fi-FI/Potilaalle/Biopankki), Finnish Red Cross Blood Service Biobank (www.veripalvelu.fi/verenluovutus/biopankkitoiminta), and Terveystalo Biobank (www.terveystalo.com/fi/Yritystietoa/Terveystalo-Biopankki/Biopankki/). All Finnish Biobanks are members of BBMRI.fi infrastructure (www.bbmri.fi). Finnish Biobank CooperativeFINBB (https://finbb.fi/) is the coordinator of BBMRI-ERIC operations in Finland. Finnish biobank data can be accessed through Fingenious® services (https://site.fingenious.fi/en/) managed by FINBB. CSC–IT Center for Science, Finland, is acknowledged for computational resources.

## Supplementary figures

**Figure S1.**
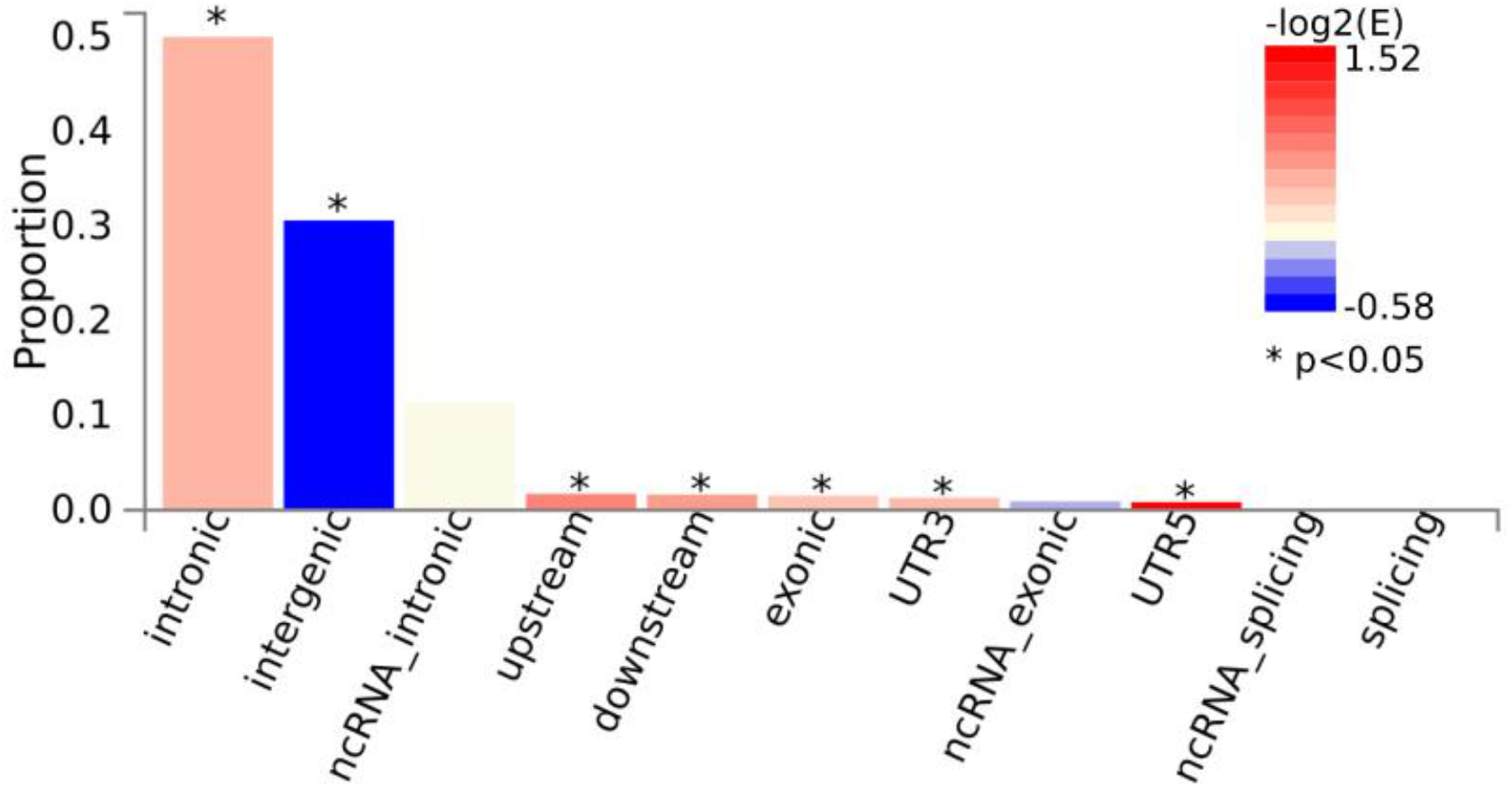
Functional annotation of the genome-wide significant variants and their LD proxies. Coloring of the bars shows -log_2_(enrichment) compared to the reference set of variants.

**Figure S2.**
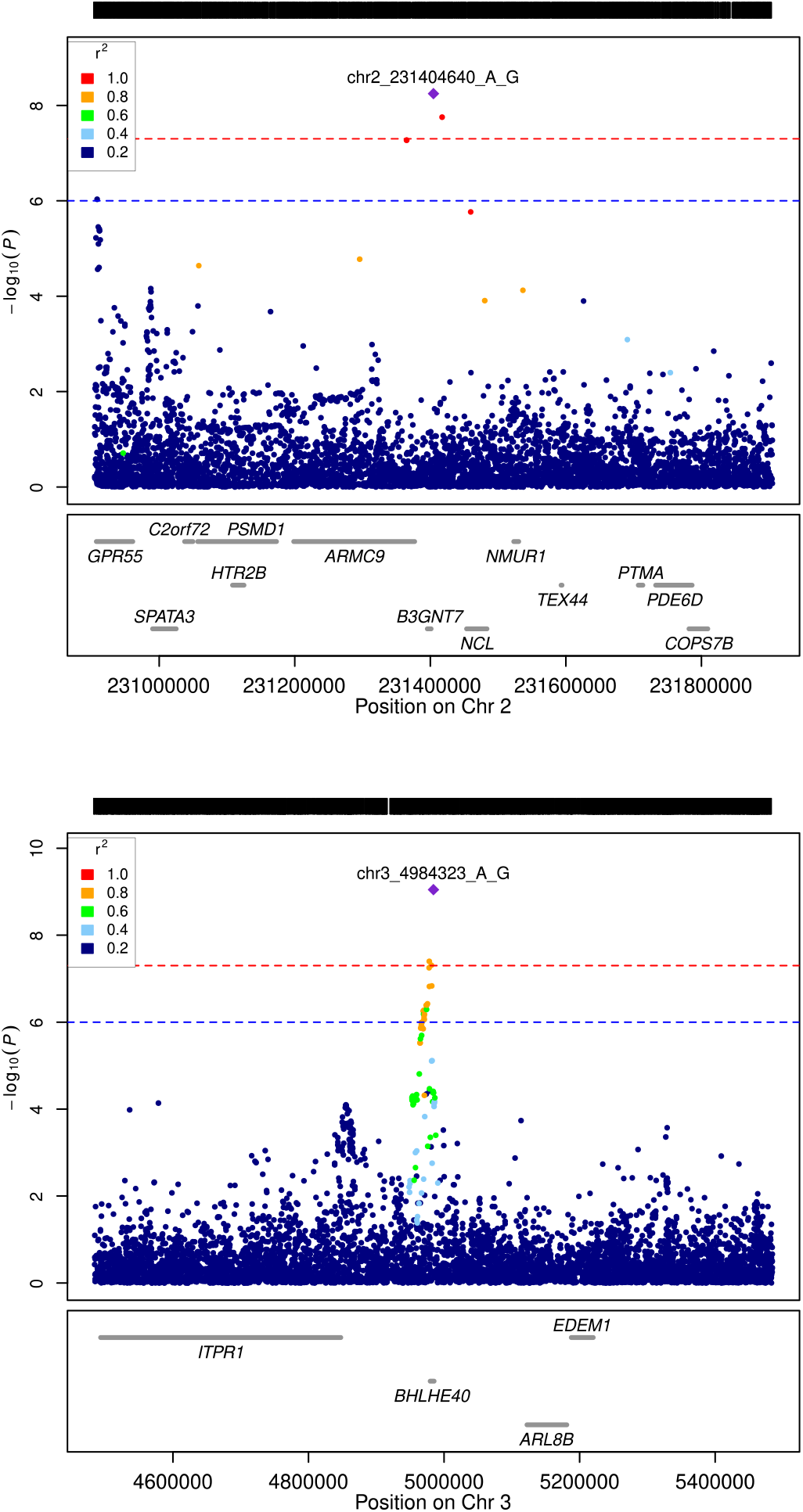
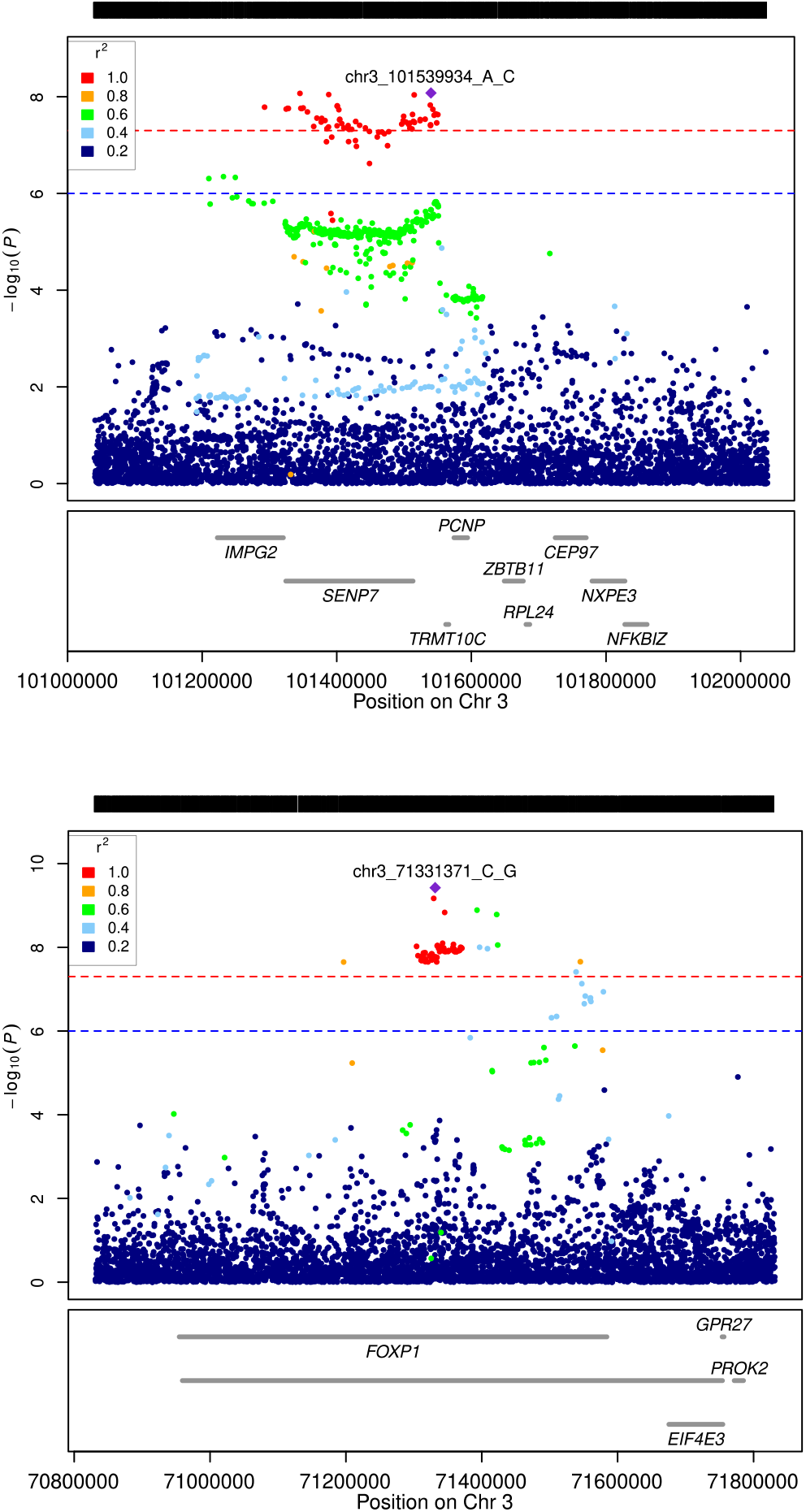
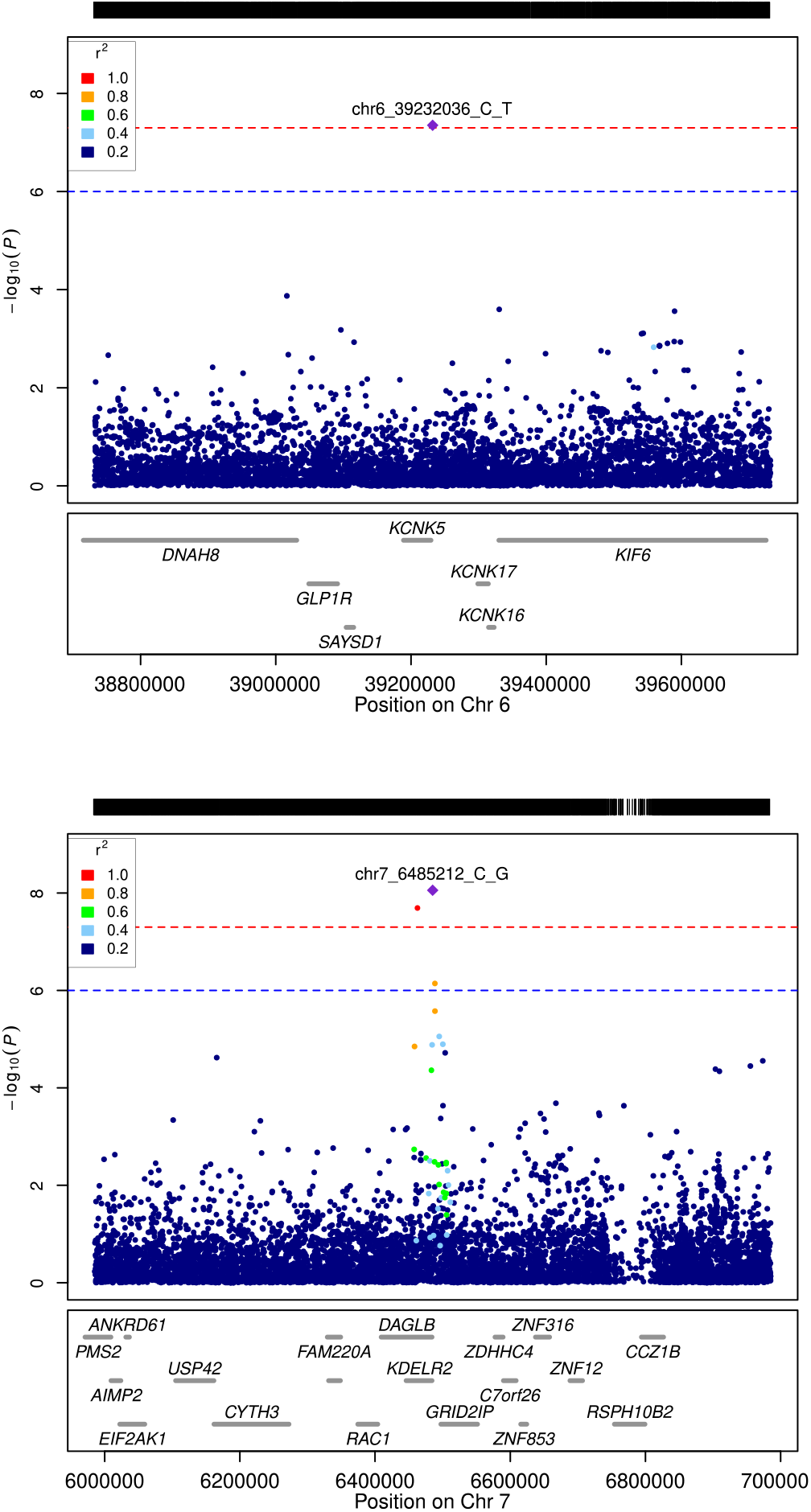
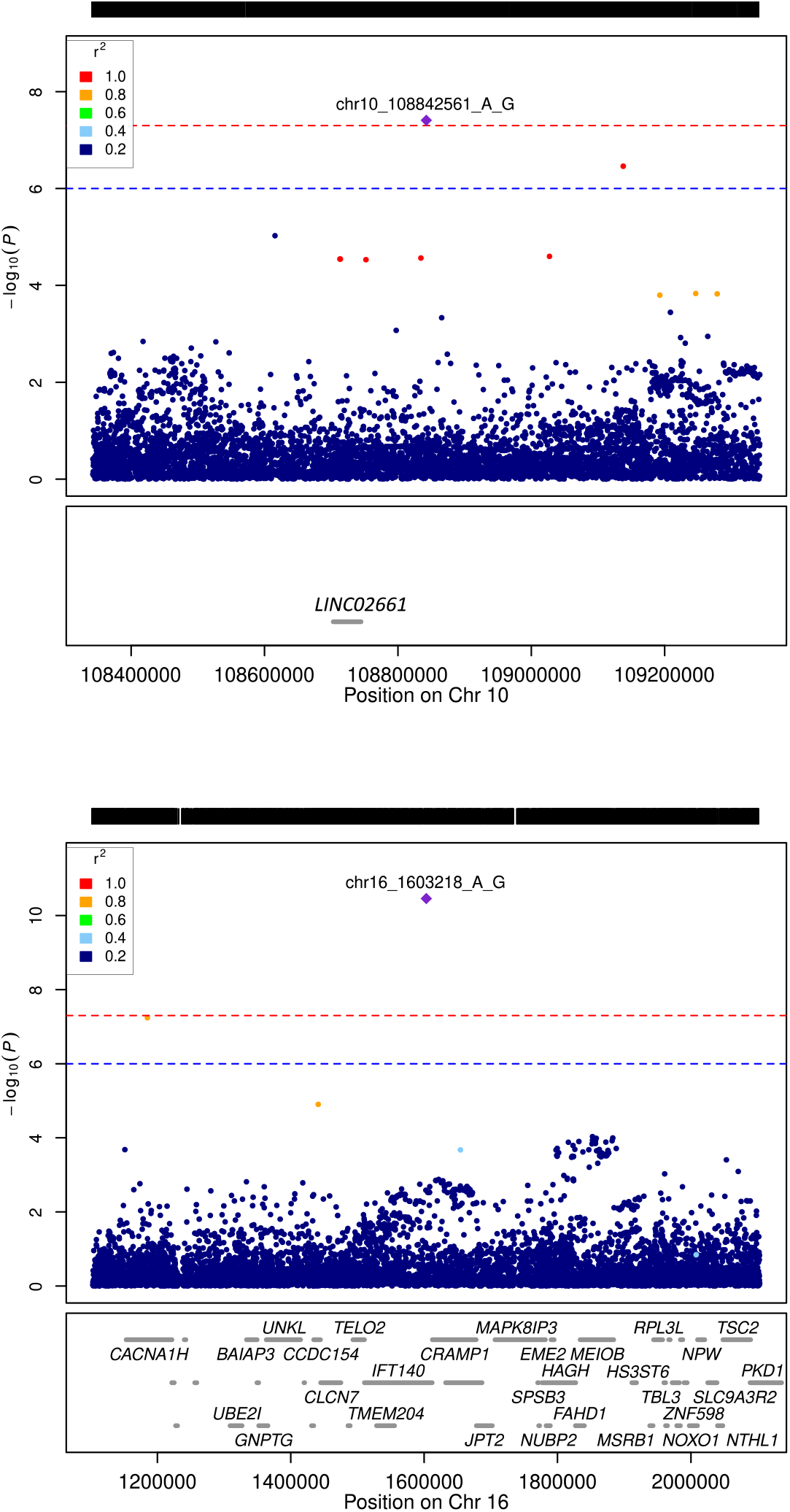
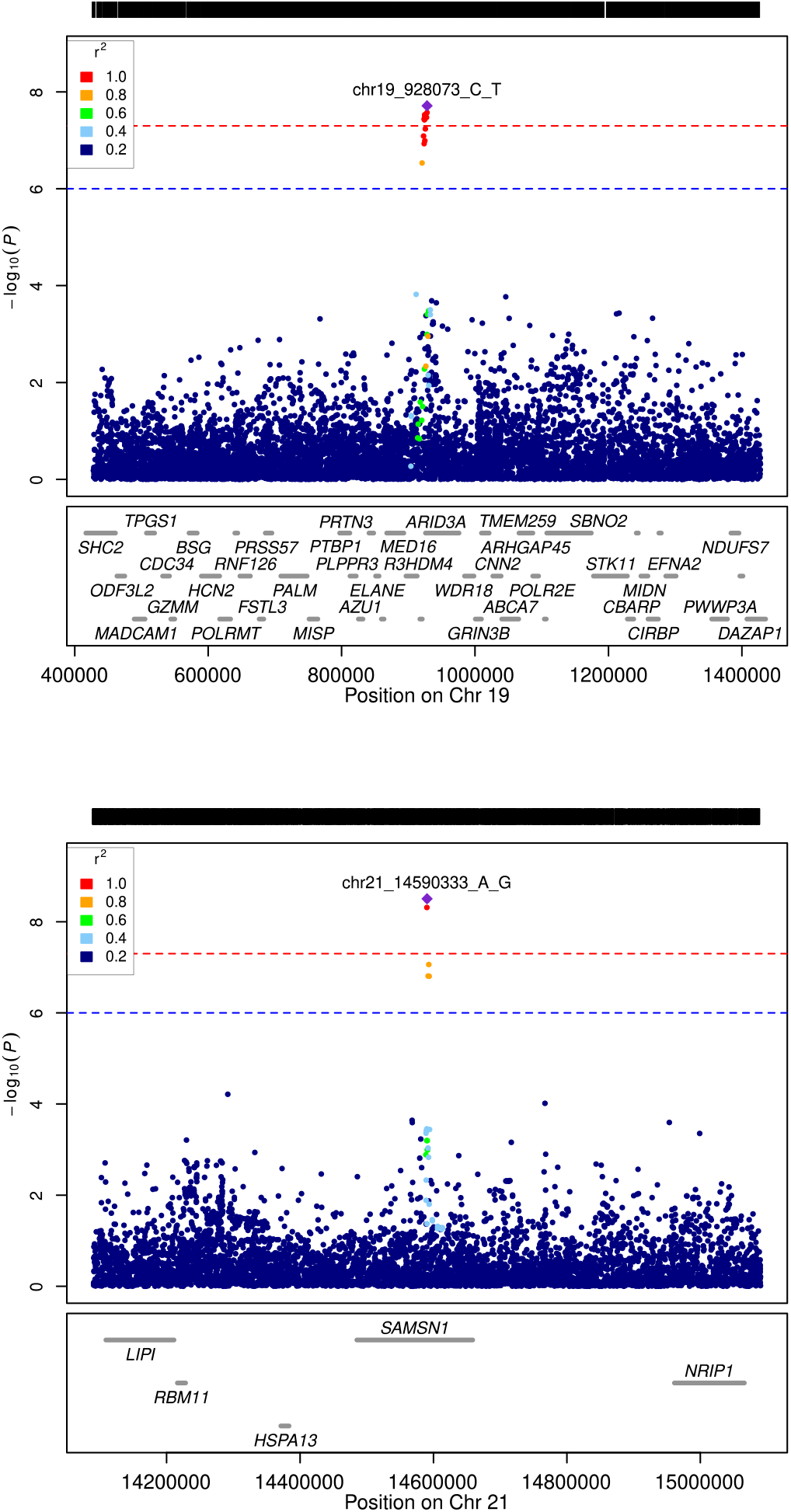
Regional association plots of the novel AD-associated loci.

**Figure S3.**
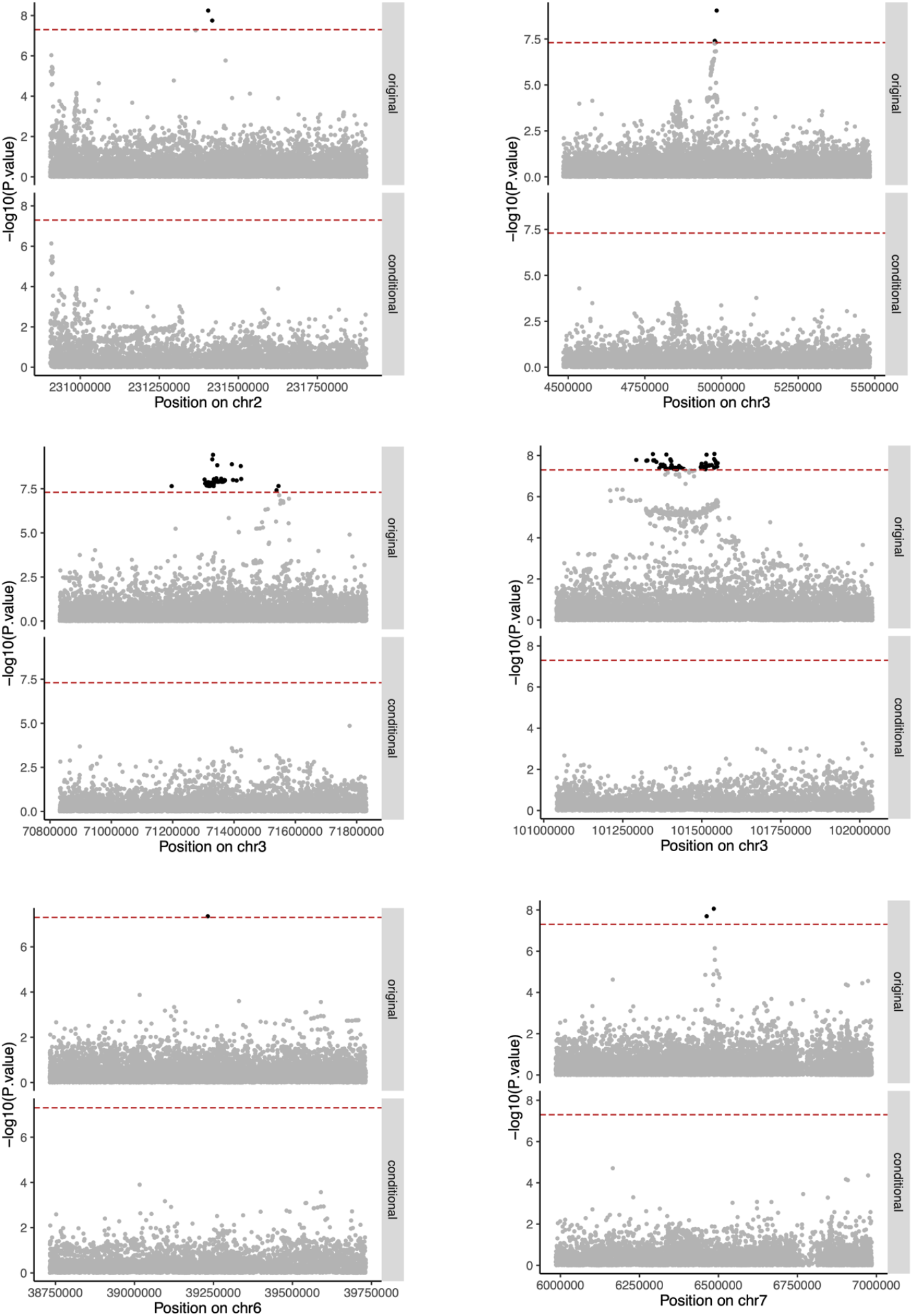
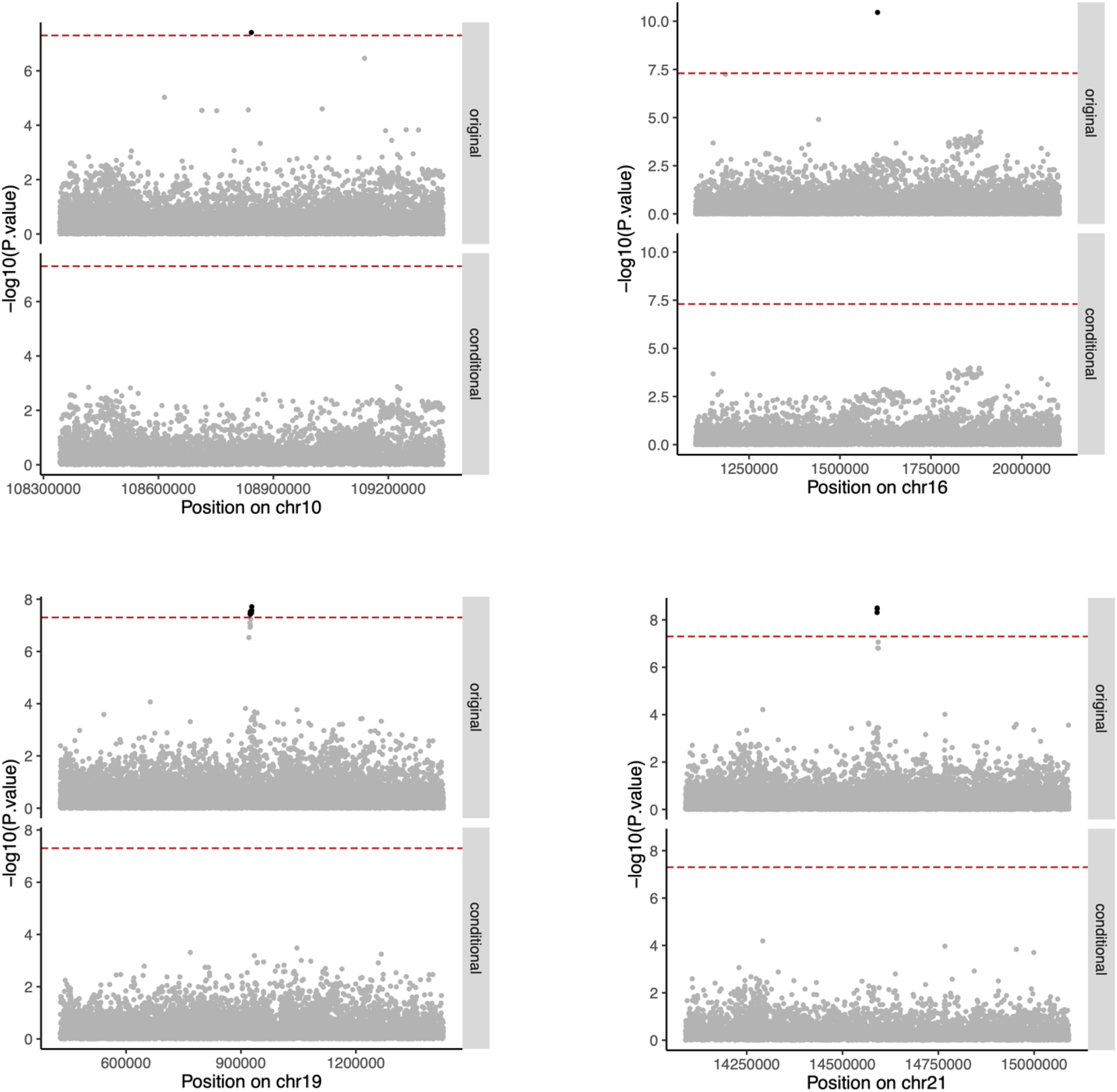
Conditional association tests of the novel AD-associated loci.

**Figure S4.**
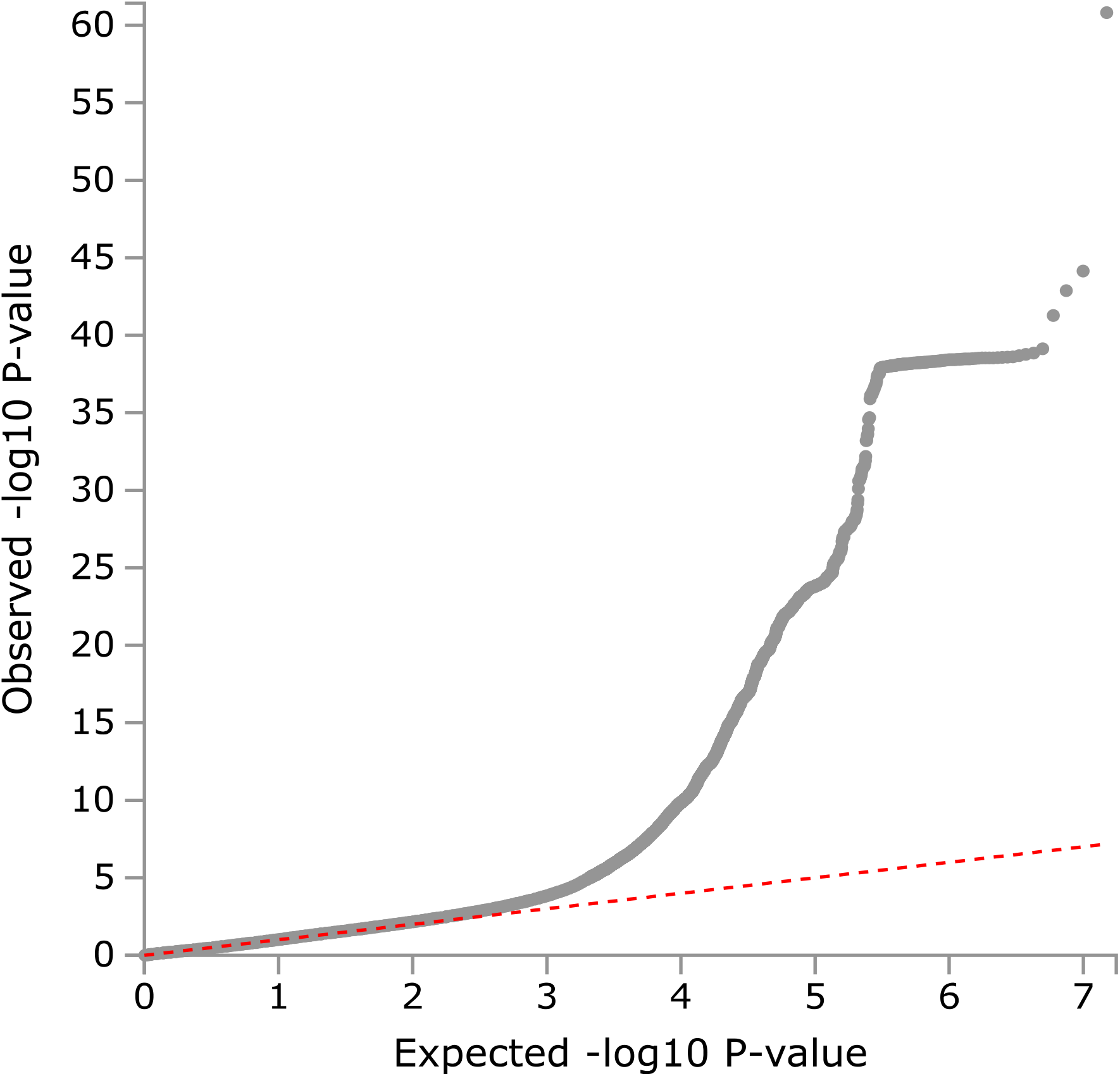
QQ plot of the genome-wide meta-analysis.

**Figure S5.**
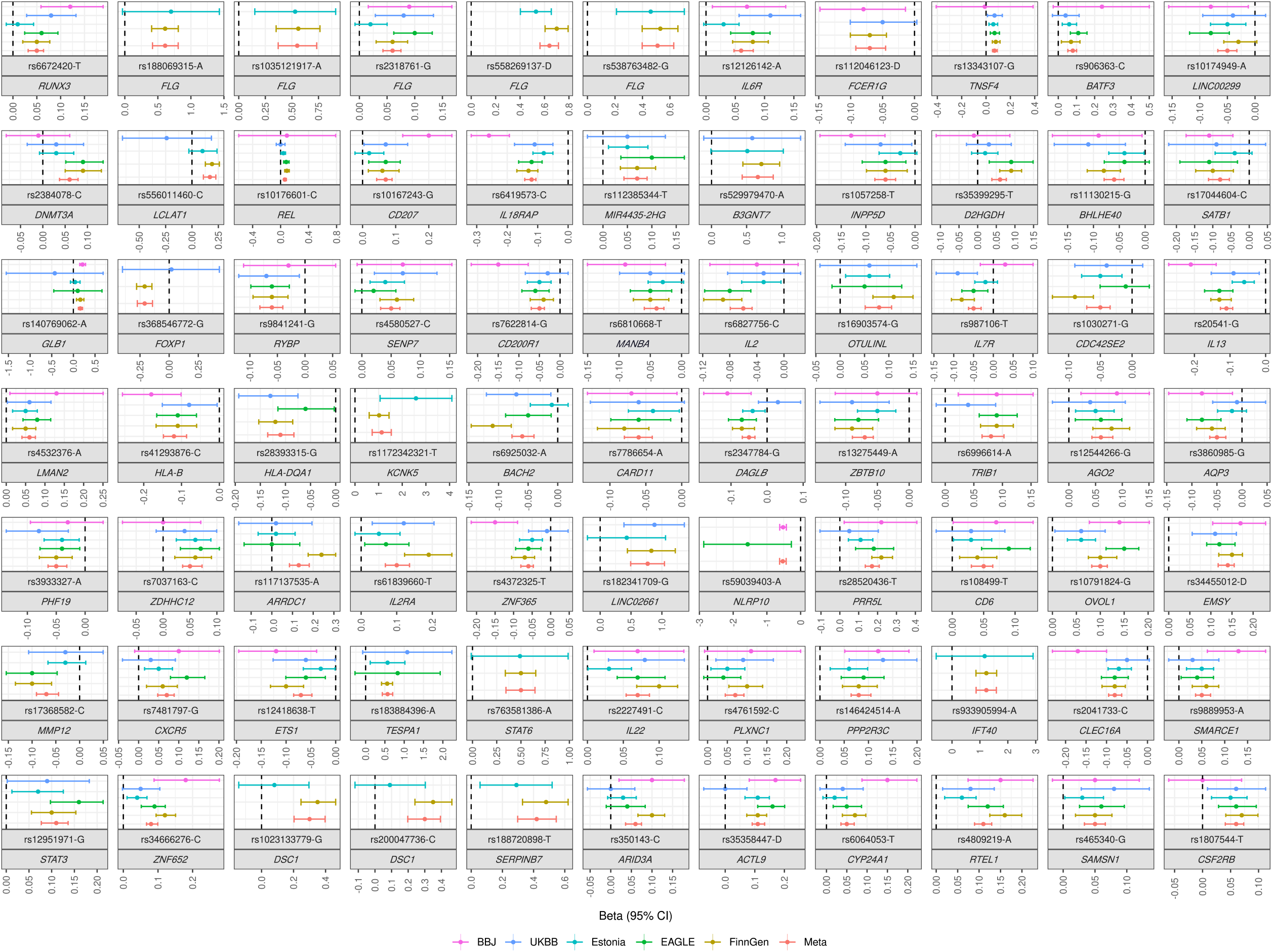
Effect estimates of the 77 genome-wide significant loci with 95% confidence intervals in the meta-analysis populations.

**Figure S6.**
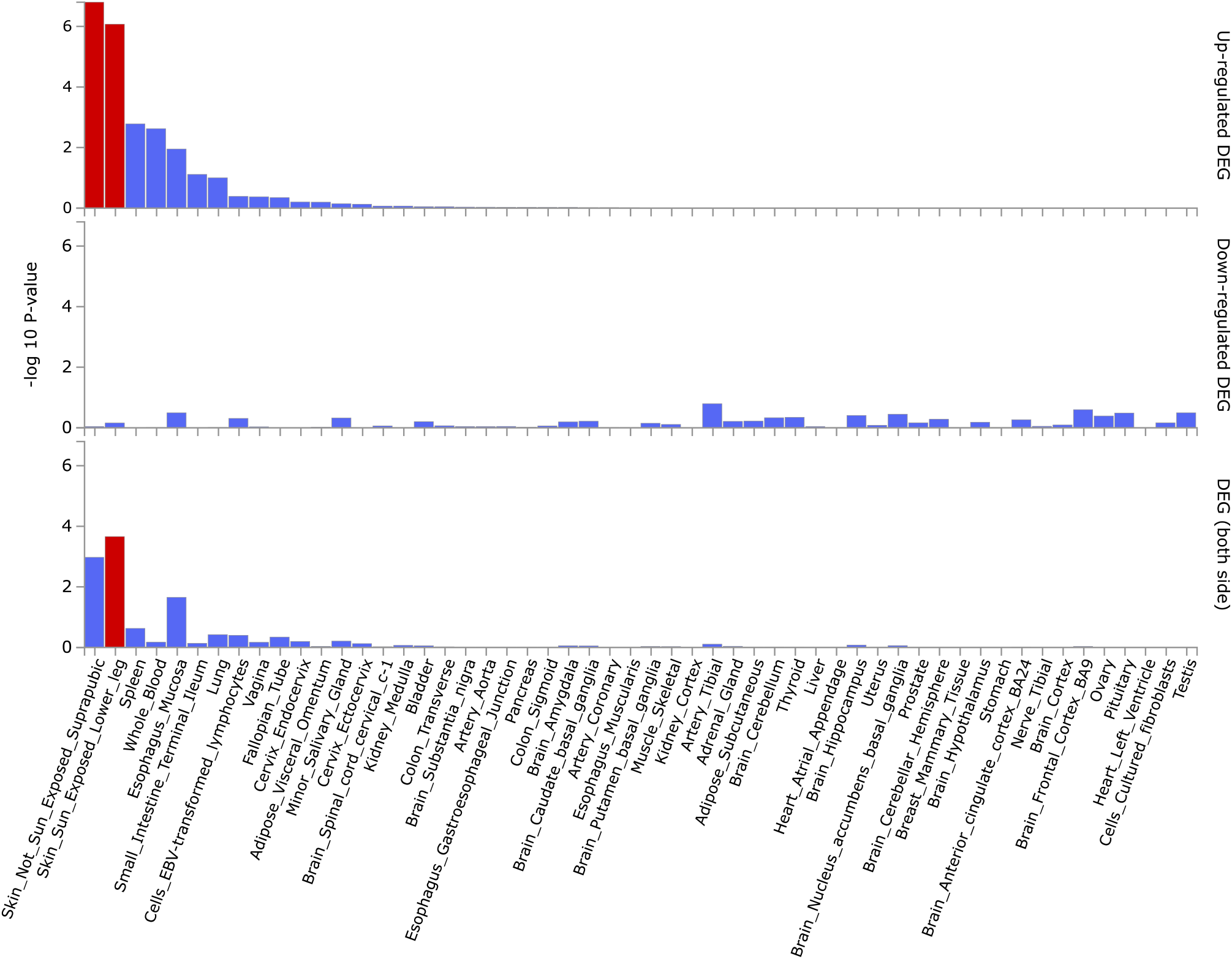
Enrichment test of the AD-associated genes for differentially expressed gene sets (DEGs) across the 54 tissue types in GTEx v8. Enrichment was significant in upregulated and two-sided DEGs in one or both of the skin tissues.

**FigureS7.**
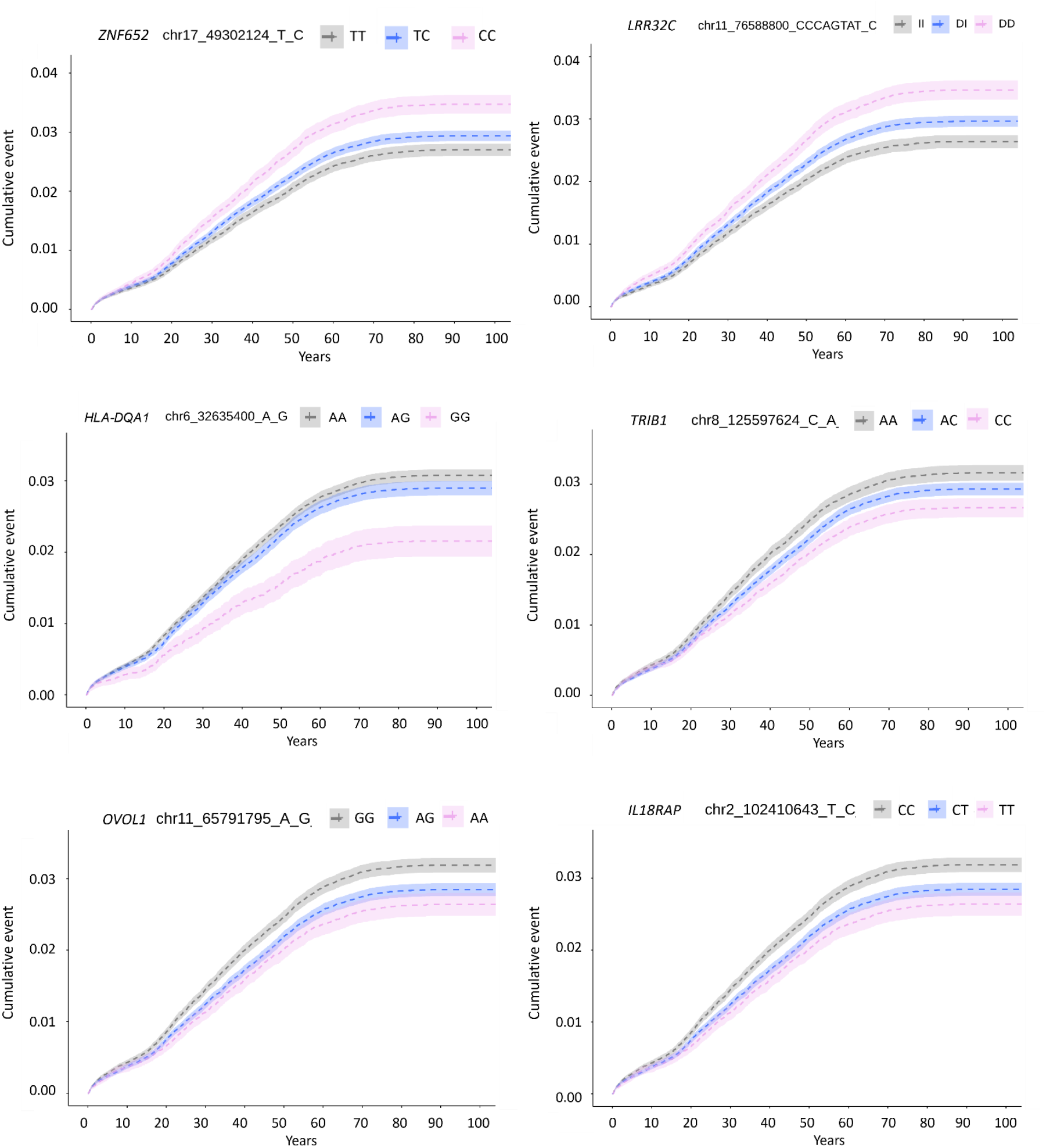
Kaplan-Meier plots of the meta-analysis loci with evidence for non-additive genetic effects.

**Figure S8.**
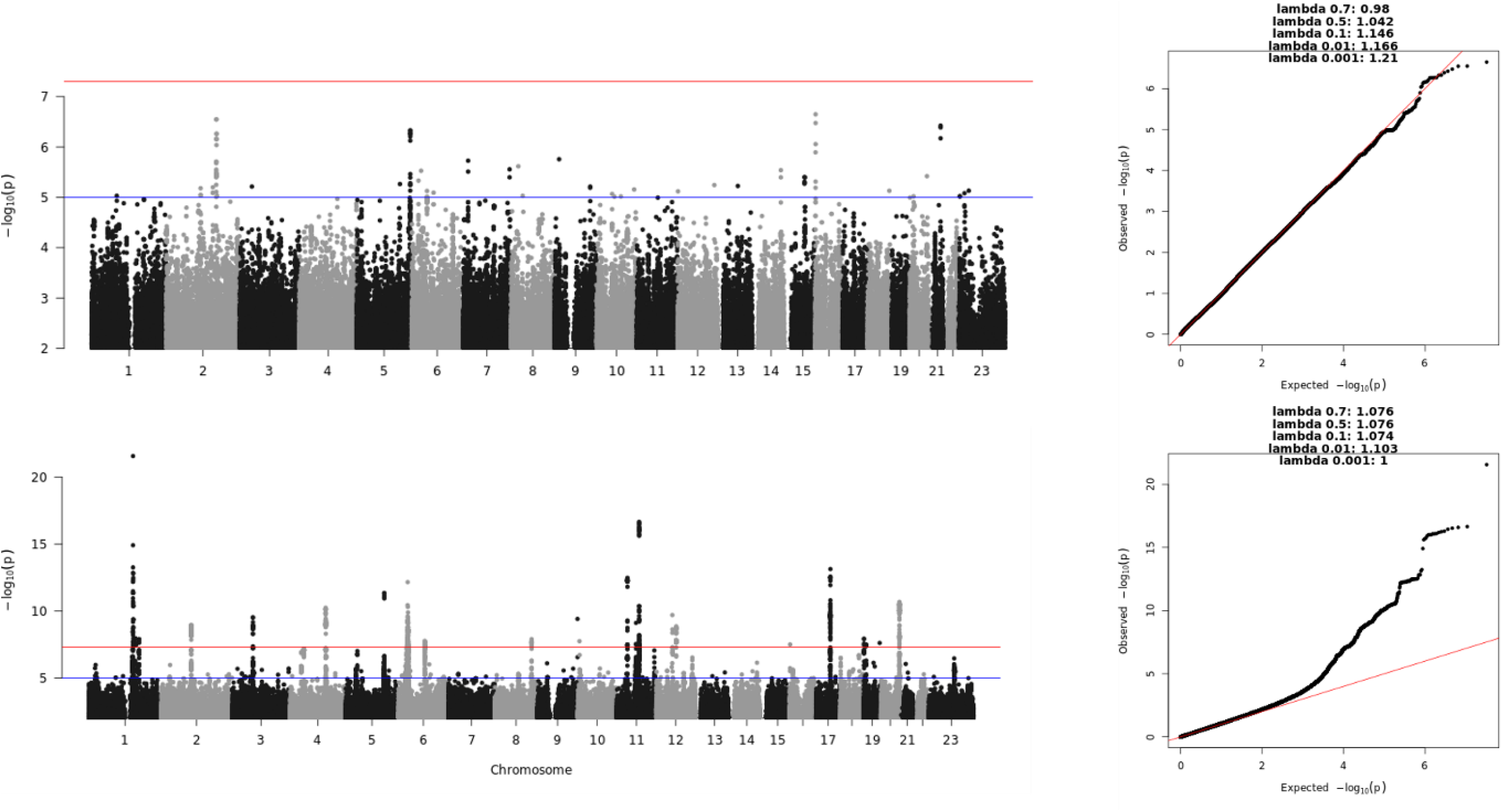
Genome-wide association analysis of early (above) and late-onset AD in the FinnGen freeze7 data.

**Figure S9.**
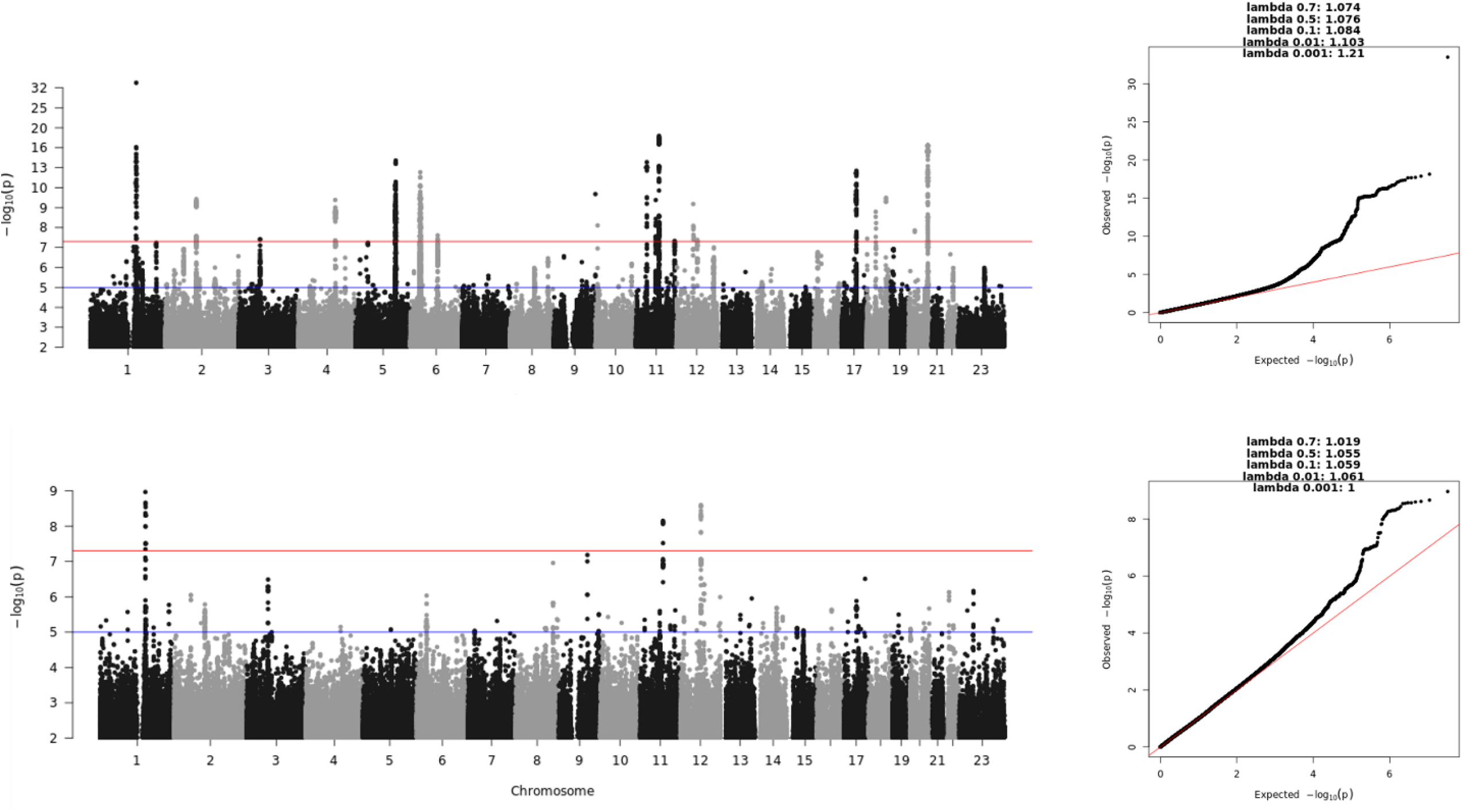
Genome-wide association analysis of mild (above) and severe AD in the FinnGen freeze7 data.

**Figure S10.**
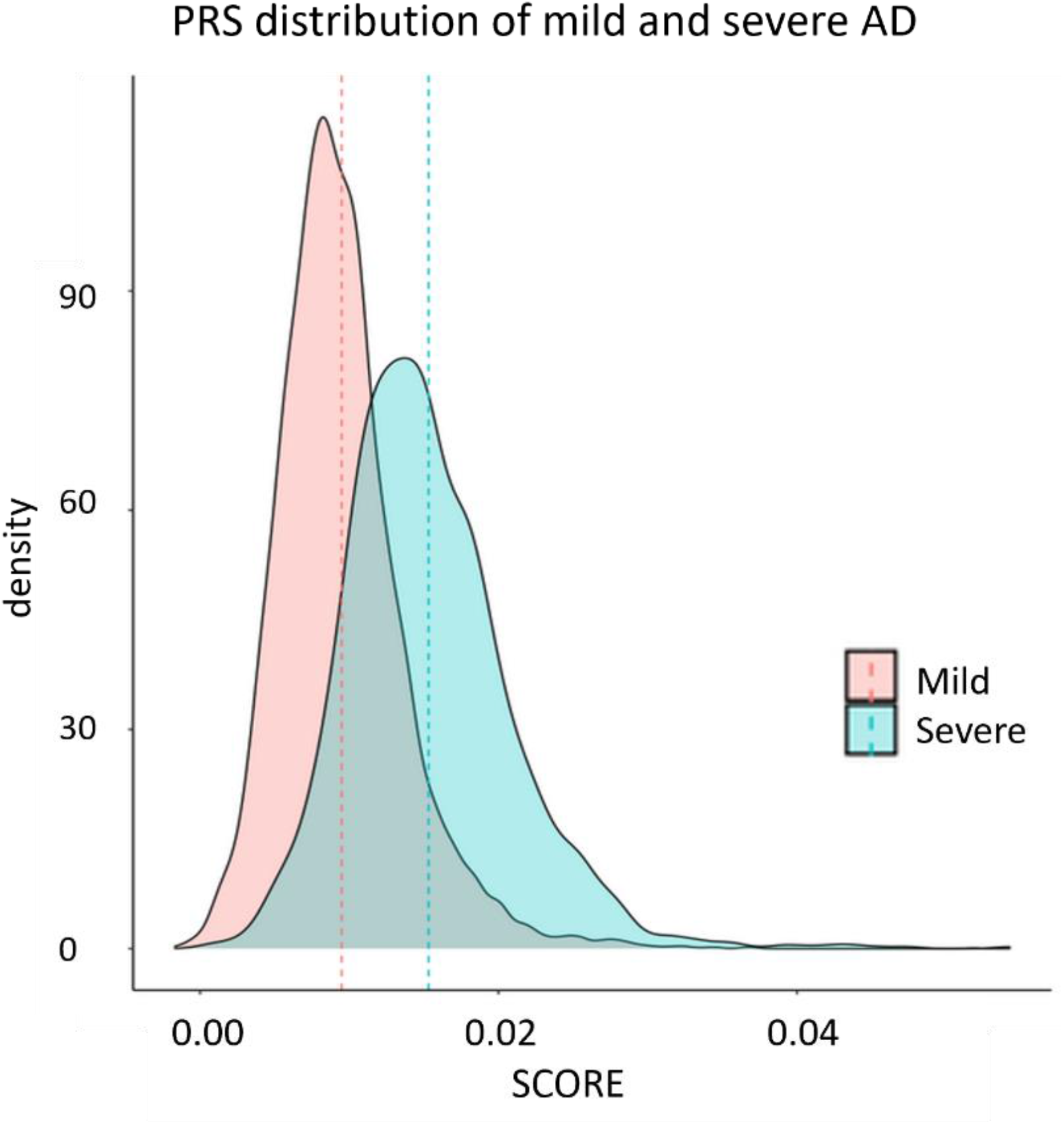
Polygenic risk scores of the mild and severe AD.

## Notes

### Competing Interest Statement

The authors have declared no competing interest.

### Author Declarations

FinnGen ethics statement with permit numbers is presented under the Methods section.

